# Nuclear genetic control of mtDNA copy number and heteroplasmy in humans

**DOI:** 10.1101/2023.01.19.23284696

**Authors:** Rahul Gupta, Masahiro Kanai, Timothy J. Durham, Kristin Tsuo, Jason G. McCoy, Patrick F. Chinnery, Konrad J. Karczewski, Sarah E. Calvo, Benjamin M. Neale, Vamsi K. Mootha

## Abstract

Human mitochondria contain a high copy number, maternally transmitted genome (mtDNA) that encodes 13 proteins required for oxidative phosphorylation. Heteroplasmy arises when multiple mtDNA variants co-exist in an individual and can exhibit complex dynamics in disease and in aging. As all proteins involved in mtDNA replication and maintenance are nuclear-encoded, heteroplasmy levels can, in principle, be under nuclear genetic control, however this has never been shown in humans. Here, we develop algorithms to quantify mtDNA copy number (mtCN) and heteroplasmy levels using blood-derived whole genome sequences from 274,832 individuals of diverse ancestry and perform GWAS to identify nuclear loci controlling these traits. After careful correction for blood cell composition, we observe that mtCN declines linearly with age and is associated with 92 independent nuclear genetic loci. We find that nearly every individual carries heteroplasmic variants that obey two key patterns: (1) heteroplasmic single nucleotide variants are somatic mutations that accumulate sharply after age 70, while (2) heteroplasmic indels are maternally transmitted as mtDNA mixtures with resulting levels influenced by 42 independent nuclear loci involved in mtDNA replication, maintenance, and novel pathways. These nuclear loci do not appear to act by mtDNA mutagenesis, but rather, likely act by conferring a replicative advantage to specific mtDNA molecules. As an illustrative example, the most common heteroplasmy we identify is a length variant carried by >50% of humans at position m.302 within a G-quadruplex known to serve as a replication switch. We find that this heteroplasmic variant exerts *cis*-acting genetic control over mtDNA abundance and is itself under *trans*-acting genetic control of nuclear loci encoding protein components of this regulatory switch. Our study showcases how nuclear haplotype can privilege the replication of specific mtDNA molecules to shape mtCN and heteroplasmy dynamics in the human population.

## INTRODUCTION

Mitochondria are ancient organelles that contain a tiny, high copy number circular genome (mitochondrial DNA, mtDNA). Sequencing of the human mtDNA in 1981 (Anderson et al., 1981) revealed that it encodes 13 core protein components of the oxidative phosphorylation system, as well 2 rRNAs and 22 tRNAs required for their expression. The remaining ∼1100 mitochondrial proteins, including all proteins required for mtDNA maintenance, replication, and transcription, are encoded by the nuclear DNA (nucDNA) and imported (Rath et al., 2020). Tissues can contain tens to thousands of copies of mtDNA per cell depending on cell type (D’Erchia et al., 2015). Variants in mtDNA can be maternally transmitted or arise somatically, and when they co-exist with wild-type molecules, lead to a state called heteroplasmy. While mtDNA maintenance is fully reliant on nucDNA-encoded proteins, a systematic understanding of how nuclear genetic variation influences variation in mtDNA abundance and heteroplasmy levels in humans is lacking.

Defects in mtDNA are associated with a spectrum of human diseases (Frazier et al., 2019). Since the first identification of pathogenic mtDNA mutations (Holt et al., 1988; Wallace et al., 1988), scores of maternally inherited syndromes have since been characterized (Ratnaike et al., 2021). Mendelian forms of mitochondrial disease producing mtDNA deletion or depletion were later identified and mapped to nuclear genes involved in mtDNA replication, maintenance, and nucleotide balance (Nishino et al., 1999; Suomalainen et al., 1995; van Goethem et al., 2001). More generally, a quantitative decline in mtDNA copy number (mtCN) and an accumulation of somatic mtDNA mutations have both long been associated with aging and age-associated disease (Ashar et al., 2017; Fazzini et al., 2021; Wanagat et al., 2001). Mutations in mtDNA accumulate in many cancers and in a small subset of tumors fulfill criteria as “drivers” of tumorigenesis (Gopal, Calvo, et al., 2018; Gopal, Kübler, et al., 2018).

Heteroplasmy dynamics are complex and presumed to be shaped by mutation, drift, and selection. The mtDNA mutation rate has been reported as 10-100x higher than the nucDNA (W. M. Brown et al., 1979; Thomas & Wilson, 1991), with the main non-coding region (control region, CR) containing three hypervariable regions thought to be mutational hotspots (Stoneking, 2000). The high copy number, elevated substitution rate, and lack of recombination have made mtDNA CR variants a valuable genetic tool in anthropology and forensics, even leading to the African mitochondrial “eve” hypothesis (Cann et al., 1987; Vigilant et al., 1991). Heteroplasmy can vary across siblings, attributed to germline bottleneck effects, and between cell types and tissues, thought to be due to random segregation and selection (Li et al., 2015; Walker et al., 2020). Mechanisms underlying heteroplasmy dynamics in humans remain obscure, though classical mouse studies identified nuclear quantitative trait loci (QTLs) controlling mtDNA segregation (Battersby et al., 2003), suggesting a role for nucDNA variation.

Here, we characterize the spectrum of mtCN and heteroplasmy across ∼300,000 individuals spanning 6 ancestry groups in UK Biobank (UKB) and AllofUs (AoU) and identify their nuclear genetic correlates. To our knowledge, this is the largest analysis of human mtDNA sequence to date. After rigorous blood cell composition corrections, we find that mtCN declines with age, is influenced by numerous nuclear genetic loci, and does not decline in most common diseases.

mtDNA heteroplasmy shows two patterns: heteroplasmic single nucleotide variants (SNVs), which tend to be somatic and accumulate with age; and heteroplasmic indels, which are more common than SNVs, occur most frequently in the non-coding region, do not vary with age, and are quantitatively inherited as mixtures of multiple alleles along the maternal lineage. These indels are present in most individuals, showing variation across the population and even across single cells from one person. For the first time, we find that many heteroplasmies are influenced by a shared nuclear genetic architecture nominating genes with established roles in mtDNA replication and maintenance as well as mitochondrial genes with no prior links to mtDNA biology. These loci are likely acting by conferring a replicative advantage to specific mtDNA sequences. For instance, the most common heteroplasmy, found in more than 50% of the population, is a length variant in the mtDNA CR, which controls mtCN (in *cis*) and itself is influenced by nuclear loci (*trans*) implicated in a mitochondrial transcription/replication switch.

## RESULTS

### Calling mtDNA copy number and variants at scale

We developed mtSwirl, a scalable pipeline for calling mtDNA variants and copy number from whole genome sequencing data (**Methods, Supplementary note 1**). We augmented a pipeline previously used to analyze mtDNA variation in gnomAD (Laricchia et al., 2022), now constructing self-reference sequences for each sample using homoplasmic and homozygous calls on the mtDNA and reference nucDNA regions of mtDNA origin (NUMTs, **Supplementary figure 1A**). mtSwirl shows improved mtDNA coverage, particularly among African haplogroups (**Supplementary figure 1B-E**), and reduced variant calls at very low heteroplasmy (**Supplementary figure 1F**), indicating reduced ancestry- and NUMT-specific mis-mapping. We observe high concordance of heteroplasmy estimates with the prior method used in gnomAD (R^2^ = 0.996 for heteroplasmy > 0.05), with homoplasmies showing allele fractions now closer to 1 suggesting reduced influence of NUMTs (**Supplementary figure 1G**, Laricchia et al., 2022). We used mtSwirl to quantify mtDNA traits across 274,832 individuals of diverse ancestry across UKB and AoU (**Supplementary figure 2, Supplementary table 1**), generating >7,800,000 mtDNA variant calls across all samples.

### Determinants of mtDNA copy number variation

We began by identifying covariates of blood mtDNA copy number (mtCN_raw_) in UKB. Our analysis highlights the strong influence of blood cell composition on mtCN_raw_ (R^2^ ∼23%, **Figure 1A**) as previously reported (Hägg et al., 2020; Hurtado-Roca et al., 2016, **Supplementary figure 3C**). We identified several additional technical covariates including time of day, month of year, and fasting duration (R^2^ ∼ 2.5%, **Figure 1A, Supplementary figures 3E-3J**). Following adjustment for all identified covariates (**Methods, Supplementary note 2, 3**), we find that corrected mtCN (which we term mtCN_corr_) was unimodal in UKB across 178,134 subjects with an average of 61.66 copies per nuclear genome (**Supplementary figure 3D**). We observed a linear decline in mtCN_corr_ with age (**Figure 1C**) of approximately 2% per decade among both males and females.

**Figure 1.**
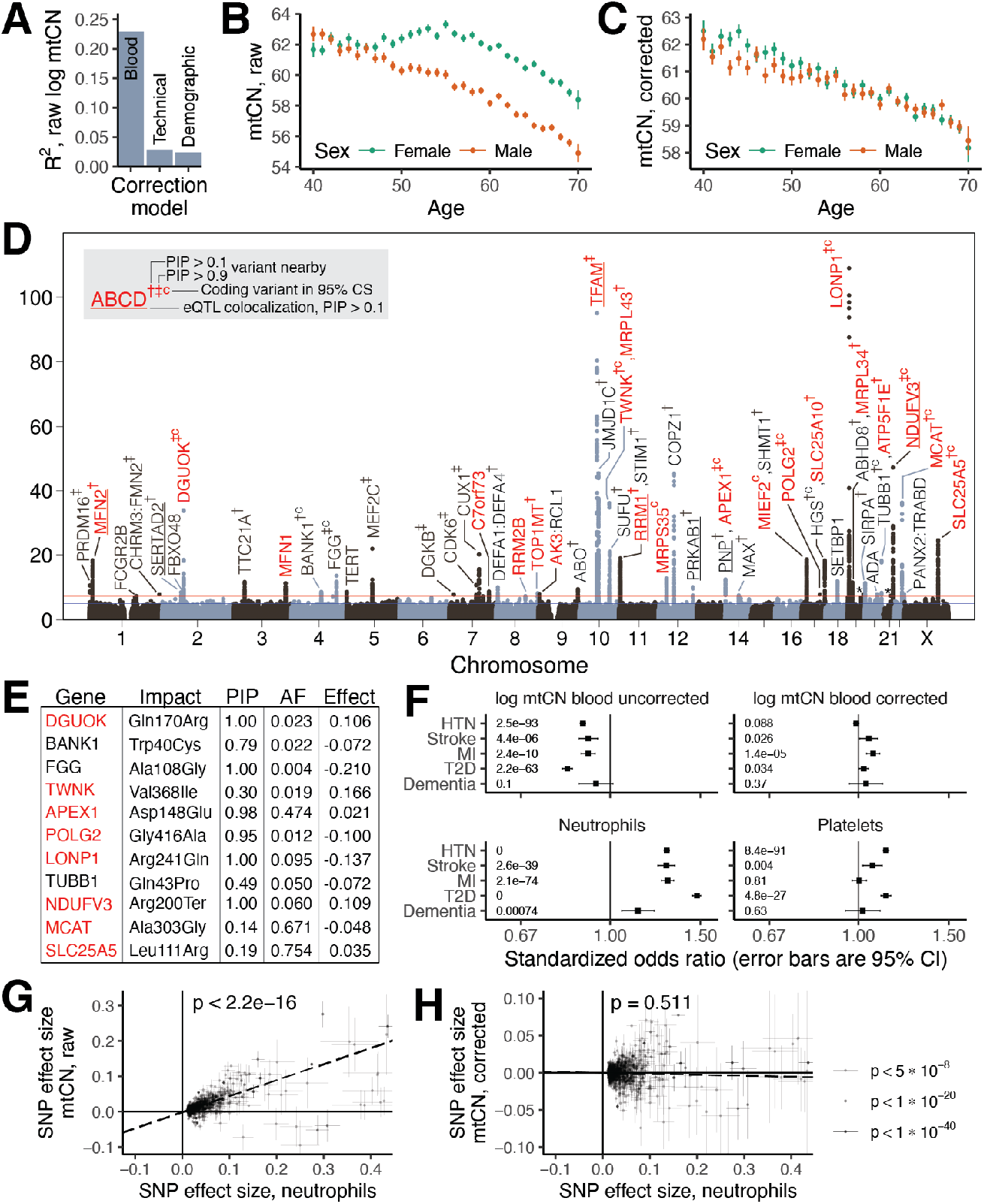
Genetic and phenotypic determinants of mtDNA copy number in UK Biobank. **A**. Variance explained in mtCN by blood composition, technical, and demographic correction models. Relationship of **B**. mtCN_raw_ and **C**. mtCN_corr_ as a function of age and genetic sex. **D**. GWAS Manhattan plot from cross-ancestry meta-analysis in UKB. Labeled genes were obtained either via fine-mapping or, if a credible set (CS) could not be constructed, mapping to the nearest gene. Red genes are mitochondrial or are implicated in mtDNA disease; † = CS variants proximal to the gene with posterior probability of inclusion (PIP) > 0.1; ‡ = CS variants with PIP > 0.9; “c” = coding variant in the CS; underline = eQTL colocalization PIP > 0.1. Asterisks above peaks on chromosome 19 and 21 correspond to GP6 and RUNX1 respectively. **E**. Table of variants in the 95% CS with PIP > 0.1 causing a protein-altering change. Red indicates mitochondria-relevant. **F**. Standardized odds ratios for log mtCN_raw_, log mtCN_corr_, and major blood composition phenotypes in predicting risk of selected common diseases in UKB. Inset numbers are p-values; error-bars are 95% CI. HTN = hypertension; MI = myocardial infarction; T2D = type 2 diabetes. Correlation between effect sizes for genome-wide significant lead SNPs detected for neutrophil count between neutrophil count and **G**. mtCN_raw_ and **H**. mtCN_corr_. Error bars represent 1SE, dotted line is weighted least squares regression line, inset corresponds to regression p-value.

We next assessed the degree to which mtCN_corr_ is under nuclear genetic control. Our GWAS identified 92 linkage disequilibrium (LD)-independent nucDNA association signals across 46 loci (**Figure 1D**) after cross-ancestry meta-analysis, with an estimated SNP-heritability of ∼4% (**Methods**). In contrast, mtDNA haplogroup explained < 0.5% of the variance in mtCN_corr_ with only a few associations of small magnitude observed (**Supplementary figure 4A, B**). 33 nuclear loci showed variants with a posterior inclusion probability (PIP) of 0.1 or greater after fine-mapping (**Methods**); 11 of these had protein-altering variants in the 95% credible set (CS) at PIP > 0.1 (**Figure 1E**) and seven showed eQTL colocalization with the assigned gene at PIP > 0.1 including *TFAM, MFN2, NDUFV3*, and *RRM1*. Seven loci contained genes implicated in disorders of mtDNA maintenance, six of which harbored variants with PIP > 0.1. Prioritized genes (**Methods**) encoded proteins that participate in the mtDNA nucleoid and replisome (*TFAM, POLG2, TWINKLE, TOP1MT, LONP1*), nucleotide metabolism (*RRM1, RRM2B, DGUOK, AK3, SLC25A5*), and mitochondrial fusion (*MFN1, MFN2*). The PNP/APEX1 locus was notable as these adjacent genes encode proteins in nucleotide metabolism and mtDNA repair, neither of which has been implicated in mtCN control. Fine-mapping implicated both genes, even identifying a missense variant in APEX1 at PIP > 0.9 (**Supplementary figure 5A**). Several additional loci included mitochondrial proteins with no prior links to mtDNA (*SLC25A10, MCAT, MIEF2, NDUFV3*). Telomerase (*TERT*) is in the vicinity of one locus, however fine-mapping did not provide additional evidence for its causality (**Supplementary table 3**).

We next tested mtCN_corr_ for heritability enrichment in genes associated with organelles or organs using stratified LD-score regression (S-LDSC, Finucane et al., 2015, 2018; Gupta et al., 2021), **Methods**). Encouragingly, the most significant organelle enrichment was seen for the mitochondrion (**Supplementary figure 4C**). Across organs, skeletal muscle and whole blood were top scoring (**Supplementary figure 4D**). Whole blood enrichment is expected given the sampling site, but skeletal muscle enrichment was unexpected and may be due to shared patterns of gene expression between blood and muscle or non-cell autonomous control of blood mtCN.

### Blood cell composition confounds prior genetic and phenotypic associations with mtCN

Although many prior studies have reported associations between low blood mtCN and common diseases (Ashar et al., 2017; Chong et al., 2022; Fazzini et al., 2021; Yang et al., 2021), we could not replicate these results using mtCN_corr_ in UKB for type 2 diabetes, myocardial infarction, stroke, hypertension, or dementia (**Figure 1F**). We tested 24 other common diseases and only observed lower mtCN in individuals with osteoarthritis (**Supplementary figure 3K**). Upon repeating this using mtCN_raw_, without adjusting for blood composition, we recovered these prior associations (**Figure 1F, Supplementary figure 3K**). Even the oft-reported elevated mtCN in females (Ding et al., 2015) appears to be largely driven by blood composition (**Figure 1B, 1C**). Our genetic analyses underscore the confounding effects of blood composition in previous work. We replicated (at p < 5*10^−5^) 70 of the 96 previously reported mtCN GWAS loci (Longchamps et al., 2021) using mtCN_corr_, with 37 at genome-wide significance (GWS) (**Methods**). However, we recover 12 additional loci from this prior study at GWS using mtCN_raw_ including loci containing *HBS1L-MYOB, C2, HLA, GSDMC*, and *CD226*, which are linked to blood cell types and inflammation (**Supplementary figure 4F**). In contrast, associations near *TFAM*, a well-known mtCN controlling gene (Ekstrand et al., 2004), strengthen by ∼40 orders of magnitude following blood composition correction. It has long been known that inflammation is associated with cardiometabolic disease (Aul et al., 2002); indeed, elevations in inflammatory blood cell indices predict elevated risk for 26/29 tested diseases in UKB (**Figure 1F, Supplementary figure 3L**). Bidirectional Mendelian randomization showed that effect sizes for GWS loci for neutrophil count were strongly positively correlated with corresponding mtCN_raw_ effect sizes (**Figure 1G**) while the converse did not convincingly hold (**Supplementary figure 4G**), suggesting that changes in blood cell composition cause mtCN_raw_ changes rather than the reverse. Importantly, neutrophil count effect sizes did not predict corresponding mtCN_corr_ effect sizes (**Figure 1H, Supplementary figure 4H**). The most parsimonious explanation for our observations is that previously reported associations between blood mtCN and common diseases are, in many cases, secondary to blood composition changes.

### Nuclear genetic control of variation in coverage across the mtDNA genome

Whole genome sequencing (WGS) yields high coverage across the 16,569 bases of the mtDNA, but it is non-uniform (**Figure 2A**). We observe a coverage dip by over 50% in the major non-coding segment of the mtDNA called the control region (CR), which contains the light strand promoter (LSP), three conserved sequence blocks (CSBs), the heavy strand origin of replication (O_H_), and the D-loop, which contains a stable third strand of DNA (7s DNA) (**Supplementary figure 6**). mtDNA replication starts with RNA primer synthesis from LSP-CSBII (red dashed arrow, **Figure 2A**). Primed mtDNA synthesis begins at CSBII, with the nascent DNA between CSBII and O_H_ forming a transient flap called the “DNA primer” (black dashed arrow, **Figure 2A**). Further replication produces the persistent 7s DNA after which replication proceeds (black solid arrow, **Figure 2A**; c.f. Falkenberg & Gustafsson, 2020). In theory, we expect the highest local WGS coverage in the persistently triple-stranded 7s DNA, lower coverage in the transiently triple-stranded “DNA primer” region, and lowest coverage in the RNA primer region. This is what we observe (**Figure 2A**).

**Figure 2.**
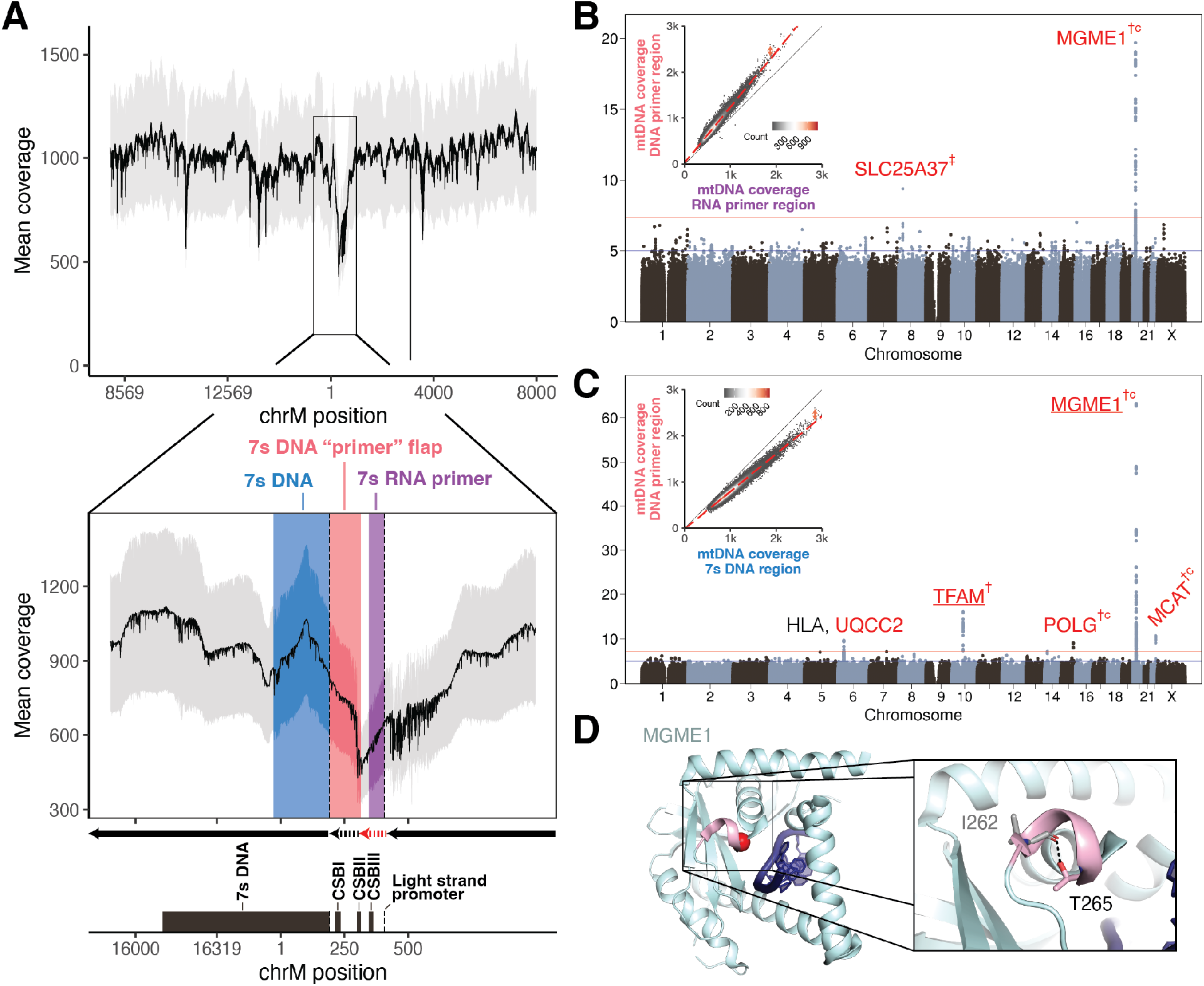
Nuclear genetic control of relative mtDNA coverage in the non-coding region. **A**. Mean per-base coverage across the mtDNA in UKB. Zoomed dropdown highlights coverage depression in the mtDNA non-coding region. Arrows correspond to stages of replication: red dashed arrow = RNA primer; black dashed arrow = transient DNA “primer” flap; black solid arrow = retained replicated DNA. Grey ribbon is +/- 1 standard deviation. CSB = conserved sequence box. **B**. GWAS Manhattan plot of the residual of the regression of mtDNA median DNA primer coverage on median RNA primer coverage. **C**. GWAS Manhattan plot of the residual of the regression of mtDNA median DNA primer coverage on median 7s DNA region coverage. Insets for **B** and **C** show 2D histograms of the correlation between the respective quantities across all individuals in UKB. Red genes are mitochondrial or are implicated in mtDNA disease; † corresponds to CS variants proximal to the gene with posterior probability of inclusion (PIP) > 0.1; ‡ corresponds to CS variants with PIP > 0.9, “c” corresponds to a missense variant in the CS; underline corresponds to eQTL colocalization PIP > 0.1. **D**. Structure of MGME1 (5ZYV) shown with bound ssDNA in dark blue, the 3_10_ helix in pink and the T265 alpha carbon as a red sphere. Inset shows the hydrogen bond between T265 and I262.

We hypothesized that genetic variation in nuclear-encoded mtDNA replication machinery might influence the persistence of replication intermediates in the CR. To quantify these intermediates, we computed the difference in coverage between these three regions across individuals in UKB (insets, **Figures 2B** and **2C, Methods**). Upon performing GWAS and cross-ancestry meta-analysis for these traits, we find that nuclear genetic variants near *MGME1* associate with the degree of coverage discordance between the RNA primer and the DNA primer (**Figure 2B**), while variants near *TFAM, POLG, MCAT, and MGME1* associate with the discordance between 7s DNA and the DNA primer (**Figure 2C**). All four genes encode mitochondrial-localized proteins, and MGME1 and POLG work in concert to resolve flap intermediates (i.e., the DNA primer) via exonuclease activity during mtDNA replication (Uhler et al., 2016). Missense variants in *POLG, MGME1*, and *MCAT* all show PIP > 0.1 after fine-mapping, and the highest PIP variant at the *MGME1* locus causes p.Thr265Ile which disrupts a hydrogen bond within a helix-forming part of the DNA binding pocket of the MGME1 exonuclease domain (**Figure 2D**), potentially impacting DNA binding. We also identify a variant causing p.Ala303Gly in *MCAT*, which has no prior connection to mtDNA maintenance and encodes a component of mitochondrial type II fatty acid synthase.

### Intermediate disease phenotypes in carriers of pathogenic mtDNA mutations

We next considered mtDNA sequence variation in UKB (**Methods**), with an initial focus on ten established pathogenic mtDNA mutations, including those associated with Leber’s hereditary optic neuropathy, mitochondrial encephalopathy, lactic acidosis, and stroke-like episodes (MELAS), and aminoglycoside-induced ototoxicity (**Figure 3**). We find that ∼1:192 individuals in UKB carry at least one of the ten variants, in agreement with a previous estimate of 1:200 (Elliott et al., 2008). A longstanding question is whether carriers of rare pathogenic mtDNA variants in the population exhibit intermediate disease phenotypes, which can now be addressed thanks to the rich phenotyping in UKB. We tested four phenotypes traditionally associated with these mtDNA variants: hemoglobin A1c (chrM:3243:A,G), triglyceride levels (chrM:3243:A,G), hearing impairment (chrM:1555:A,G, chrM:3243:A,G, chrM:7445:A,G), and visual impairment (chrM:3460:G,A, chrM:11778:G,A, chrM:14484:T,C, chrM:14459:G,A) (M. D. Brown et al., 2000; Rydzanicz et al., 2011; Sharma et al., 2021; Shoffner et al., 1995). Individuals carrying the chrM:3243:A,G variant show elevated hemoglobin A1c, elevated triglycerides, and hearing and vision impairment (**Figure 3, Methods**). The other tested mtDNA variants were not associated with deviations in these phenotypes.

**Figure 3.**
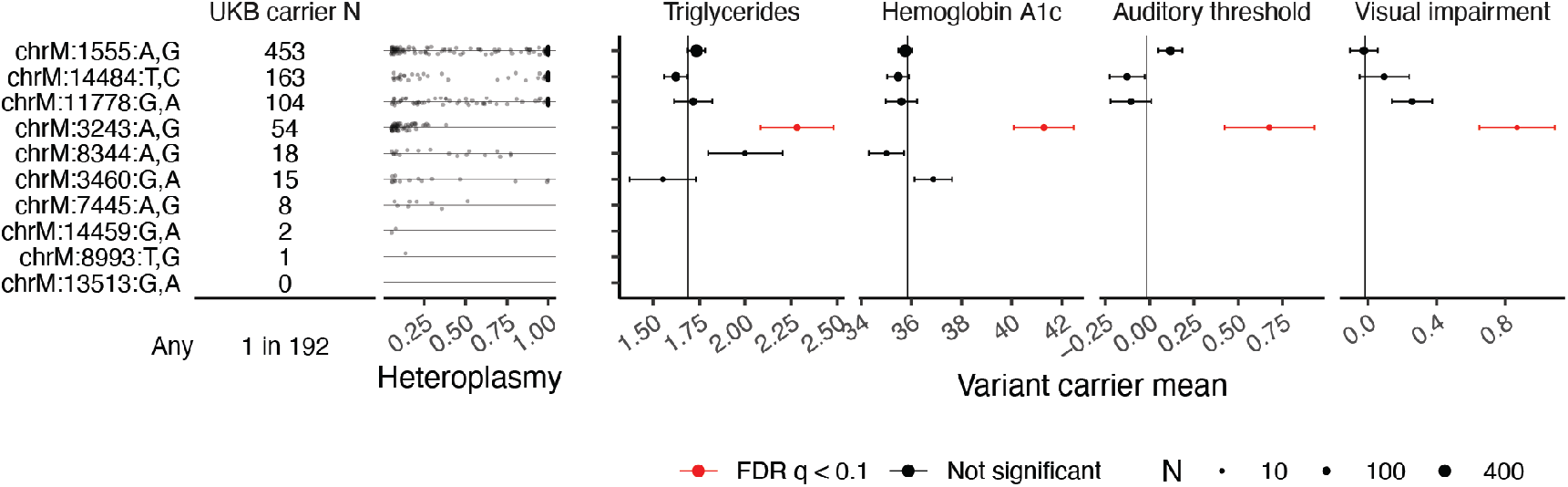
Evidence of intermediate phenotypes among carriers of the MELAS variant in UKB. Table shows carrier frequencies for 10 known pathogenic mutations in UKB, including chrM:3243:A,G (pathogenic for MELAS), with heteroplasmy distributions plotted as jittered points. Panels show mean Hemoglobin A1c, triglyceride levels, auditory threshold (via speech recognition threshold test), and visual impairment (via vision test measured as logMAR) among mtDNA pathogenic variant carriers. Only points corresponding to more than 10 measurements are shown. Vertical lines represent per-trait means among individuals with none of the 10 pathogenic mutations detected.

### Spectrum of mtDNA sequene variation across 253,583 individuals

Our analysis across UKB and AoU yields the largest database of mtDNA SNVs and indels to date (**Figure 4A**). Consistent with prior gnomAD analysis (Laricchia et al., 2022), we find that the number of homoplasmies per individual is closely related to haplogroup, with haplogroup H (closest to GRCh38 reference) showing the fewest and haplogroup L0 showing the most (**Supplementary figure 7A**). Heteroplasmy distributions were consistent between UKB and AoU (**Figure 4B, Supplementary figure 7D, 7H**), and most individuals carried 0-1 heteroplasmic SNVs and 0-2 heteroplasmic indels (**Supplementary figure 7E**). The hypervariable regions of the mtDNA, found within the non-coding CR, contain an elevated heteroplasmic SNV rate and a vast predominance of heteroplasmic indel variants (**Figure 4A**). Heteroplasmic indels primarily arise near poly-C stretches (e.g., chrM:302, chrM:567, chrM:955, chrM:16182) in the non-protein-coding mtDNA, while coding mtDNA shows a low indel rate despite the presence of many poly-C tracts (**Figure 4A**), consistent with negative selection. We tested the most common heteroplasmies in UKB for associations with risk of 29 common diseases (**Methods**) and found little or no evidence of association, though sample sizes were limited (**Supplementary figure 7F**).

**Figure 4.**
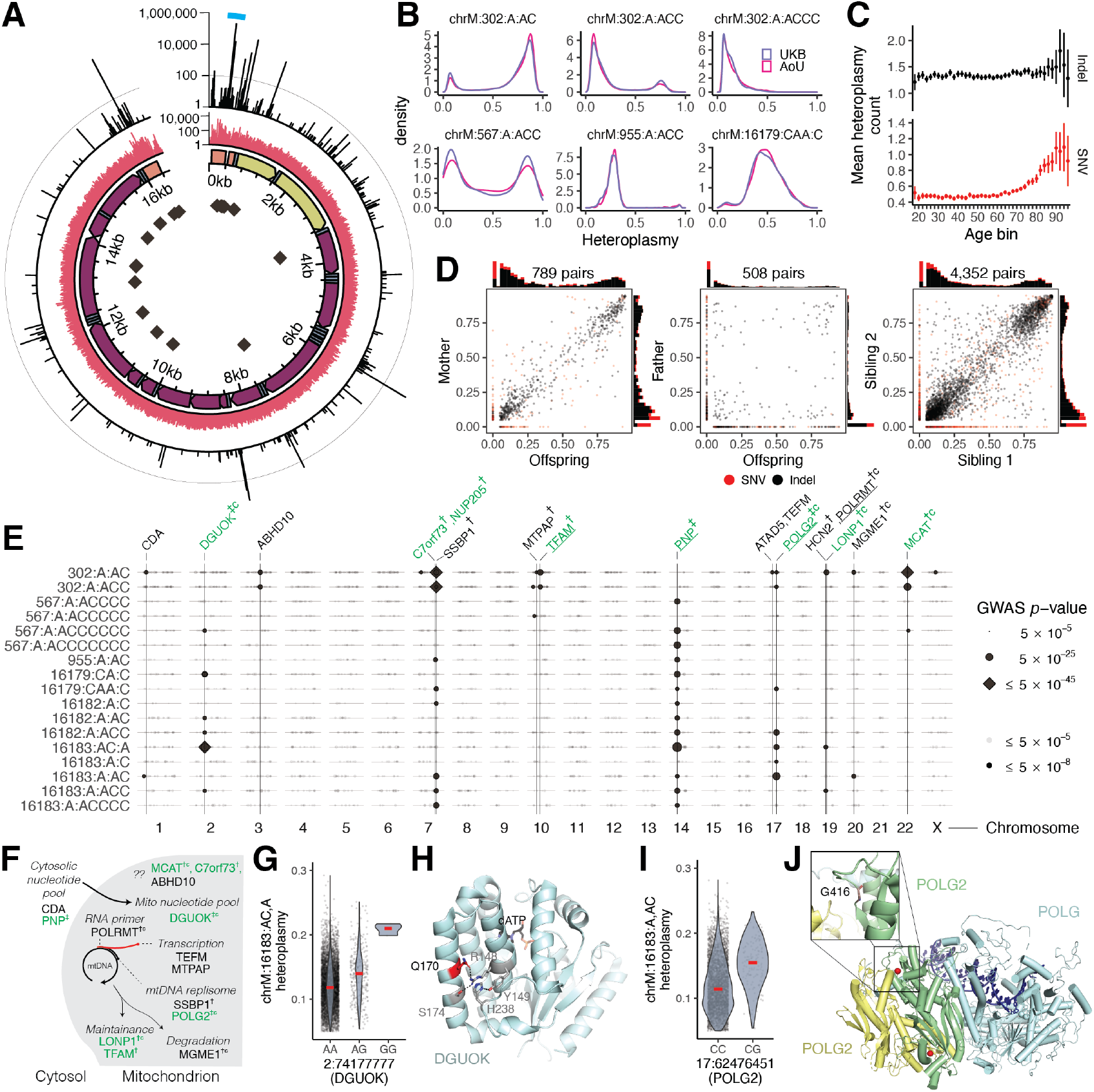
Pervasive nuclear genetic control over the most common mitochondrial DNA heteroplasmies. **A**. mtDNA heteroplasmies passing QC in UKB and AoU. Data tracks show, starting from the inside: positions of poly-C tracts; mtDNA genomic annotations (orange = HVR, yellow = rRNA genes; blue = tRNA genes; purple = coding genes); counts of heteroplasmic SNVs (red); counts of heteroplasmic indels (black). Teal arc corresponds to region highlighted in Figure 5. Light line in outermost track is a reference line at 100. **B**. Selected heteroplasmy distributions across UKB and AoU in individuals carrying the allele. **C**. Mean count of heteroplasmies per individual across age groups in AoU. Error bars are 1SE. **D**. Relationship between heteroplasmy levels in mother-offspring (left), father-offspring (middle), and sibling-sibling (right) for all heteroplasmies found in >5 individuals. **E**. GWAS lead SNPs from all common heteroplasmies with genome-wide significant signals. Point size corresponds to lead SNP p-value; dark points are genome-wide significant. Vertical lines correspond to SNPs near genes of interest and/or loci found across multiple mtDNA variants. Green corresponds to genes nominated for mtCN, † = CS variants with PIP > 0.1; ‡ = CS variants with PIP > 0.9, “c” = coding variant in CS; underline = eQTL colocalization PIP > 0.1. **F**. mtDNA dynamics pathway showing genes highlighted in heteroplasmy GWAS. **G**. chrM:16183:AC,A heteroplasmy as a function of lead SNP genotype in DGUOK. **H**. Structure of DGUOK (2OCP) with amino acid Q170 in red and nearby residues participating in hydrogen bonds or stacking interaction in pink. dATP shown as black sticks. **I**. chrM:16183:A,AC heteroplasmy as a function of lead SNP genotype in POLG2. **J**. Structure of polymerase gamma enzyme (4ZTU) with POLG in light blue and POLG2 subunits in green and yellow. Bound DNA is in dark blue and the POLG2 residue G416 is shown as red spheres. In panels G and I, red lines correspond to medians.

### mtDNA SNVs and indels exhibit distinct modes of transmission and age accrual

We next investigated the patterns of transmission and age-dependence for mtDNA heteroplasmies. For analysis of age, we focused on AoU given the broader age range of participants (20-90 versus 40-70 for UKB). While heteroplasmic SNVs tend to accumulate with age (particularly after age 70), this was not the case for indel heteroplasmies (**Figure 4C**). Using siblings and parent-offspring pairs in UKB (**Methods**), we find that nearly all heteroplasmic indels are quantitatively maternally transmitted and shared between siblings, while most heteroplasmic SNVs are not (**Figure 4D**). The maternal transmission and stability across age leads us to conclude that most indel heteroplasmies are inherited as mixtures; in contrast, for heteroplasmic SNVs, the typical lack of transmission and accumulation with age strongly suggests that they typically arise via somatic mutagenesis. In contrast to prior reports, no variants showed evidence of paternal transmission (**Figure 4D**). Transitions were far more frequent than transversions and showed a sharp increase in frequency in older age, consistent with the somatic mtDNA mutational spectrum seen in aging brains (Kennedy et al., 2013). Curiously, we observed a decline in transversion heteroplasmies in older individuals (**Supplementary figure 7F**).

### Nuclear genome GWAS for mtDNA heteroplasmy

We next sought to determine the extent to which mtDNA heteroplasmy is influenced by nuclear genetic loci. To our knowledge, nuclear genetic loci influencing individual mtDNA heteroplasmies in humans have never been reported. Given that most common heteroplasmies showed maternal transmission (**Supplementary figure 7H**), we restricted to individuals carrying each heteroplasmy and performed GWAS with the heteroplasmy level as a quantitative trait (**Figure 4B, Supplementary figure 7I**).

We identified 42 LD-independent associations across 39 heteroplasmies after cross-ancestry meta-analysis of our UKB GWAS. Our results revealed a shared nuclear genetic architecture for heteroplasmies across mtDNA sites, with nine of 20 unique nuclear loci associated with >1 heteroplasmic variant (**Figure 4E, Supplementary figure 9A**). Cross-mtDNA heterogeneity was also observed: chrM:302:A,AC and chrM:302:A,ACC appeared most associated with loci near *SSBP1, TFAM, LONP1*, and *MCAT*, while the other heteroplasmies were most strongly associated with loci containing *DGUOK, PNP*, and *POLG2*. While many genes implicated in heteroplasmy control were also identified in our mtCN GWAS, others were not (e.g., *TEFM, POLRMT, MTPAP, SSBP1, ABHD10*; **Figure 4E**). Many associated loci were near genes with established roles in mtDNA replication and maintenance (**Figure 4F**), with missense variants identified within the 95% credible set in *DGUOK, LONP1, POLRMT, MGME1*, and *POLG2* and eQTL colocalization PIP > 0.1 seen for *POLRMT, POLG2*, and *TFAM*. Of the novel hits, we highlight a locus containing *C7orf73* (**Figure 4E, Supplementary figure 9F**), which encodes a protein recently linked to Complex IV (Sang et al., 2022), suggesting a moonlighting role for this short protein in mtDNA maintenance.

Zooming in, we see strong effect sizes from PIP > 0.9 variants in or near genes related to nucleotide metabolism (*PNP, DGUOK*) and DNA replication (*POLG2*). The likely causal variant for *PNP* (PIP 1, **Supplementary figure 9G**) is in an intron of *PNP* and colocalizes with a strong negative cross-tissue eQTL (multi-tissue p ∼ 0; colocalization PIP 1; **Supplementary figure 9H-I**; Aguet et al., 2020) this gene is not yet linked to mtDNA disease but performs an analogous reaction to *TYMP* (an mtDNA disease gene) on purines. The likely causal variant for DGUOK (PIP 0.99, **Figure 4G**) results in a p.Gln170Arg missense change within the kinase domain, potentially disrupting the tertiary structure of the protein as this glutamine side chain participates in a number of hydrogen bonds and stacking interactions (**Figure 4H**). The putative causal variant for *POLG2* (PIP 1, **Figure 4I**) results in p.Gly416Ala within a predicted anticodon binding domain. This amino acid is highly conserved (**Supplementary figure 9J**) and the mutation impacts a loop near the *POLG2* homodimer surface (**Figure 4J**). These examples highlight variants impacting proteins and producing a large impact on the levels of specific heteroplasmic mtDNA variants.

To test if heteroplasmy-associated nuclear loci act via mtDNA mutagenesis, we repeated our GWAS re-coding heteroplasmy traits as “case/control”, where for each mtDNA variant, cases showed detectable heteroplasmy and controls did not. We observed little signal (**Supplementary figure 9B**), arguing against a mutagenic origin influenced by nucDNA variation and supporting the notion that maternal transmission determines the presence of each tested heteroplasmy, while nuclear variation can influence the subsequent relative heteroplasmic fraction.

We took several steps to validate our genetic findings. We performed a replication analysis in AoU across 96,698 diverse individuals and observed high concordance between cross-ancestry meta-analysis effect sizes in UKB and AoU (R^2^ = 0.79, **Supplementary figure 9C**) with limited attenuation (as expected with Winner’s curse, c.f. Lohmueller et al., 2003). We investigated potential technical and biological confounders, observing little correlation between these variables and heteroplasmies (**Supplementary figure 8A-E, Supplementary note 2**). We explicitly tested the robustness of our results to the contaminating effects of NUMTs (**Supplementary note 5**), finding that GWAS effect sizes were not sensitive to mtDNA coverage as would be expected for NUMT-derived signals (**Supplementary figure 8J-M**). Additionally, we found strong correlations between UKB meta-analysis effect sizes and those from individual ancestry groups in AoU despite small N (R^2^ = 0.49-0.78 **Supplementary figure 9D**), reducing the likelihood of confounding by recent polymorphic NUMTs. We tested all GWAS hits for LD R^2^ > 0.1 with variants within 20kb windows of 4,736 reference and polymorphic NUMTs, finding only one concerning locus – among UKB EUR, the *SSBP1* locus had LD R^2^ = ∼1 with variants in a reference NUMT. Importantly, this locus remained significant for chrM:302:A,AC among AFR in AoU despite AFR showing much lower LD with NUMT variants (**Supplementary figure 9K**).

### Pervasive length variation in CSBII across individuals and within single cells

The “length heteroplasmy” at chrM:302, located within the CSBII region of the mtDNA CR (**Figure 5A**), is the most common heteroplasmic site we observed in UKB and occurs within a regulatory motif for mtDNA replication (Wanrooij et al., 2010). Though the reference genome corresponds to G_m_AG_7_ (nomenclature indicates the length of the poly-G stretch on the GRCh38 opposite strand, **Figure 5A**), we frequently observe individuals harboring G_m_AG_8_ (chrM:302:A,AC), G_m_AG_9_ (chrM:302:A,ACC), and G_m_AG_10_ (chrM:302:A,ACCC). Quantitatively, the levels of these heteroplasmies are shared between siblings (**Figure 5B**), indicating maternal transmission of mixtures of multiple mtDNA haplotypes.

**Figure 5.**
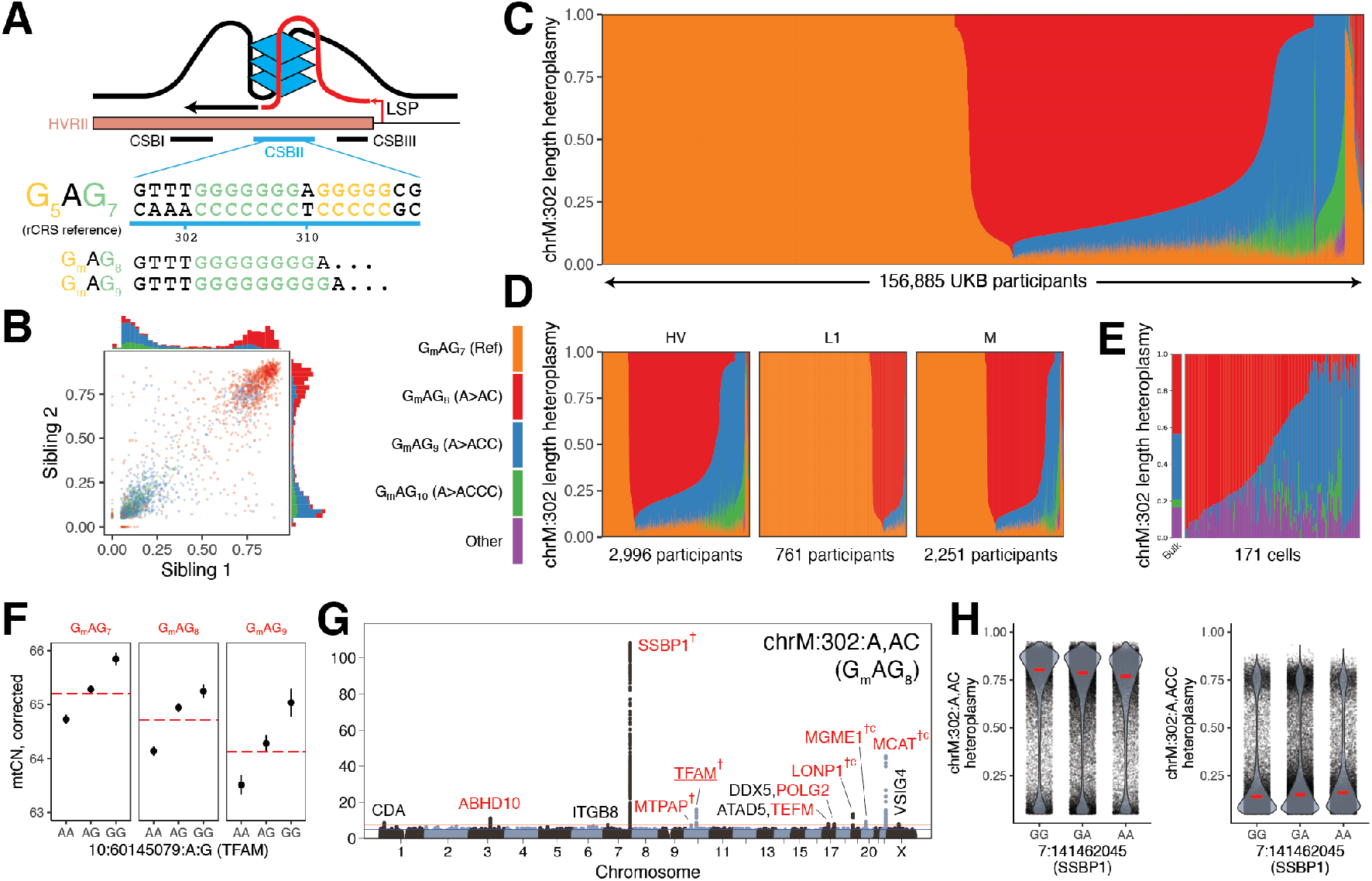
chrM:302 length heteroplasmies are inherited maternally as mixtures, co-exist in single cells, and are under the influence of the nuclear genome. **A**. Scheme showing chrM:302 region inside CSBII responsible for forming a G-quadruplex structure along with length heteroplasmy G_m_AG_n_ nomenclature. **B**. Sibling-sibling transmission of length heteroplasmies at chrM:302. **C**. Length heteroplasmy composition across all UKB individuals. **D**. Length heteroplasmy composition in UKB in select mtDNA haplogroups. **E**. Length heteroplasmy composition across 171 single cells in whole blood. Each vertical bar corresponds to a single individual (C, D) or cell (E). Colors for panels B-E correspond to legend between panels B and D. **F**. Effect of length of major allele at chrM:302 (red line) and TFAM fine-mapped variant (black dot) on mtCN. Error bars are 1SE. **G**. Case-only mtDNA heteroplasmy GWAS Manhattan plot for chrM:302:A,AC. Red genes are mitochondrial or are implicated in mtDNA disease; † corresponds to CS variants proximal to the gene with PIP > 0.1; “c” corresponds to coding variant in CS; underline corresponds to eQTL colocalization PIP > 0.1. **H**. chrM:302 length heteroplasmies as a function of highest PIP SNP genotype in SSBP1 locus. Red line corresponds to per-nuclear-genotype median heteroplasmy.

Most of the 156,885 individuals assessed in UKB harbor a mixture of these length heteroplasmies (**Figure 5C**), with individuals from different haplogroups showing different distributions (**Figure 5D**). The observed quantitative maternal transmission of heteroplasmy implies that mtDNA mixtures exist in individual cells, and we indeed find mtDNA mixtures at chrM:302 in 171 single cells from one individual (**Figure 5E**) by re-analyzing single-cell mtDNA ATAC-seq data (**Methods**).

We find multiple lines of evidence linking mtDNA replication and length variation at chrM:302. Longer alleles at this site are associated with declining mtCN_corr_ with an effect size comparable to the *TFAM* locus (**Figure 5F**, PIP ∼ 1). GWAS for chrM:302:A,AC, the most common length heteroplasmy, nominated several genes relevant for mtDNA replication and nucleotide balance not identified in other heteroplasmy GWAS (*CDA, MTPAP, TFAM, TEFM, LONP1, MCAT*; **Figure 4E, 5G**). mtCN and chrM:302:A,AC heteroplasmy even show colocalization at the two most significant mtCN loci: 10:60145079:A,G (a *TFAM* 5’ UTR variant) and 19:5711930:C,T (a *LONP1* missense variant) both show a PIP ∼ 1 for mtCN and have PIP > 0.3 for chrM:302:A,AC. It is notable that prior studies have suggested that length variation at the chrM:302 site serves as a “rheostat” for mtDNA replication versus transcription. The G-quadruplex at CSBII (**Figure 5A**) is a tertiary structure formed by the DNA and the nascent RNA primer which promotes DNA replication by blocking RNA polymerase progression (Wanrooij et al., 2010, 2012). *In vitro* studies have suggested that CSBII G-quadruplex strength is a function of chrM:302 allele, influencing the degree to which RNA transcription switches to DNA synthesis (**Figure 5A**, Tan et al., 2016). For the first time we now report that nuclear variants in genes related to the mtDNA replisome can favor one length heteroplasmy over another – for example, variants near SSBP1 favor chrM:302:A,ACC (**Figure 5H**). Taken together, our results suggest that nuclear genetic variation can influence the replication efficiency of mtDNA molecules based on chrM:302 allele.

## DISCUSSION

mtDNA heteroplasmy dynamics are highly complex, shaped by random drift and selection that in principle can operate at the level of mtDNA, mitochondria, or cells. Given that all protein machinery for mtDNA replication and maintenance is encoded by the nucDNA, it has long been theorized that the nuclear haplotype could influence mtDNA heteroplasmy. Classical mouse genetics revealed the existence of nuclear QTLs that could influence heteroplasmic mtDNA transmission (Battersby et al., 2003), though specific mechanisms and relevance to humans have been lacking. Here, for the first time, by leveraging whole genome sequencing across two large biobanks, we report pervasive nuclear genetic control of mtDNA abundance and heteroplasmy variation in humans. Many of these nuclear QTLs involve the machinery responsible for mtDNA maintenance, which likely act directly on mtDNA by altering the relative replication efficiency of mtDNA molecules based on their sequence, while several others correspond to genes never before linked to mtDNA biology. High statistical resolution allows us to gain detailed molecular insights into the mechanisms underlying an entire battery of mito-nuclear interactions, with implications for human disease, physiology, and evolution.

Our ability to dissect the genetic architecture of mtCN and heteroplasmy was possible both because of the statistical power afforded by the scale of large biobanks and because of careful attention given to technical and biological confounders. We analyzed mtDNA sequences across 274,832 individuals of diverse ancestries from two biobanks, generating the largest collection of mtDNA traits to date. We were particularly attentive to the technical challenges of contamination by mtDNA pseudogenes in the nuclear genome (NUMTs, **Supplementary Note 5, 6**). We explicitly tested many potential confounders of mtDNA traits, finding that correction of mtCN for blood cell composition had a profound effect on the observed association landscape. Many previously reported associations between blood mtCN and cardiometabolic traits (Ashar et al., 2017; Fazzini et al., 2021) disappear or reverse direction after adjustment for blood cell composition (**Figure 1F**). Our corrections reduce and even eliminate GWAS hits near genes suspiciously related to blood cell composition and inflammation (e.g., HLA, HBS1L) seen in recent studies (Longchamps et al., 2021). Our data suggest that, in many cases, an inflammatory state in cardiometabolic disease influences blood cell composition, driving the previously observed decline in mtCN.

The resulting GWAS of mtCN_corr_ and mtDNA heteroplasmies provide new insights into mtDNA maintenance. The nuclear loci we identify, including those with fine-mapped missense variation (e.g., *MGME1, POLG, POLG2, DGUOK, LONP1*), are enriched for roles in the mtDNA nucleoid, mtDNA replication, and nucleotide balance, rather than pathways previously implicated in heteroplasmy maintenance in model organisms such as mitophagy or stress response (Gitschlag et al., 2016; Lin et al., 2016). We show how population-level genetic analysis can produce detailed, mechanistic insights into mtDNA replication: GWAS of the relative mtDNA coverage in the 7*S* DNA “primer” highlights missense variants in both *MGME1* and *POLG*, whose products have exonuclease activity that can resolve this “flap” intermediate. We observe notable differences in the genetic architecture of mtCN_corr_ and heteroplasmy, producing additional insights: while *TFAM, LONP1, DGUOK*, and *PNP* are associated with both, the former two (encoding components of the mtDNA nucleoid) were the most significant associations for mtCN_corr_, while the latter two (involved in nucleotide balance) were among the strongest associations across heteroplasmies. QTLs corresponding to *TWNK* were only identified for mtCN_corr_ while associations near *SSBP1, TEFM*, and *POLRMT* were specific to heteroplasmy, suggesting that genetic variation in different mtDNA replication genes can have effects specific to mtCN or heteroplasmy. We identify many loci with no prior links to mtDNA biology, such as *C7orf73, MCAT, ABHD10, NDUFV3, CDA*, and *ADA*, proposing new roles for their protein products. The *PNP* gene product represents an excellent candidate gene for unsolved mtDNA deletion syndromes as it performs an analogous function to *TYMP* for purines and is notable for its association with the levels of 13 length heteroplasmy variants at three mtDNA sites.

A striking finding from our work is that nearly everyone harbors heteroplasmic mtDNA variants obeying two key, previously unappreciated, principles: (i) heteroplasmic SNVs are typically somatic, accrue with age sharply after age 70, and tend to be transitions, while (ii) heteroplasmic indels are found in >60% of individuals, do not accrue with age, and are usually inherited as mixtures within the same maternal lineage. Consistent with prior work (Stoneking, 2000), heteroplasmic SNVs tend to occur more in the mtDNA hypervariable regions, but most heteroplasmies detected here are inherited indels. Most heteroplasmic indels appear to occur next to poly-C stretches in the non-protein coding mtDNA; heteroplasmic indel rates are orders of magnitude lower next to poly-C stretches in coding regions, suggesting negative selection in these regions. Strikingly, for any given common indel, we find that maternal heteroplasmy levels quantitatively predict offspring heteroplasmy levels, suggesting neutral transmission. We show for the first time that these heteroplasmy levels are also under nuclear genetic control, with associated loci enriched for genes involved in mtDNA biology and nucleotide balance. These loci are similar across heteroplasmies at multiple mtDNA sites, suggesting a shared genetic architecture.

Our identified nuclear QTLs for mtDNA length heteroplasmies could, in principle, operate by one of two mechanisms: (1) the associated nuclear variants are “mutagenic” and impair mtDNA copying fidelity resulting in somatic indels due to slippage in poly-C tracts (Marchington et al., 1997), or (2) these nuclear variants confer an mtDNA replicative advantage to maternally inherited mtDNA molecules carrying certain length variants. Our data favors the latter. Case/control GWAS showed very little signal compared to case-only analysis; in concert with the observed maternal transmission this strongly suggests that the identified nuclear QTLs modify existing indel heteroplasmy levels rather than acting via mutagenesis, likely by altering the replicative efficiency of the mtDNA molecules carrying different alleles. Variants near *POLG2*, but notably not *POLG*, were associated with heteroplasmy; *POLG* is the active subunit of mtDNA polymerase in which mutations produce a “mutator” phenotype (Trifunovic et al., 2004), while *POLG2* is the accessory subunit relevant for processivity (Lim et al., 1999; Longley et al., 2006).

Our work provides insight into mechanisms by which the nuclear haplotype can confer a replicative advantage to specific mtDNA variants. This is perhaps best illustrated by length heteroplasmy at chrM:302. This heteroplasmy occurs within the G-quadruplex in CSBII in the mtDNA noncoding region, which can induce switching from transcription to replication by blocking transcription progression. Prior *in vitro* studies have shown that the chrM:302 length polymorphism impacts the strength of this G-quadruplex hence modifying the transcription/replication switch (Agaronyan et al., 2015; Tan et al., 2016). We find that mixtures of mtDNA with different chrM:302 length variants are maternally inherited in more than half of the population. Once inherited, we show that chrM:302 alleles influence mtDNA abundance (acting in *cis*), and we find that the resulting heteroplasmy levels are influenced in *trans* by nuclear QTLs (e.g., *SSBP1, POLG2, TEFM*) whose proteins directly operate this regulatory switch (Tan et al., 2016). In sum, our results suggest that the associated nuclear variants alter chrM:302 heteroplasmy by influencing factors that interact with the chrM:302 G-quadruplex, thus privileging the replication of mtDNA templates carrying a particular chrM:302 genotype. Recent experiments in embryonic stem cells led to speculation that CSBII length variants may contribute to mtDNA reversion after mitochondrial replacement therapy (MRT) (Kang et al., 2016) due to replicative advantage of carryover mtDNA from the intending mother – our nuclear genetic association results may provide insight into nuclear genetic control of this reversion.

An open question is why mtDNA heteroplasmy is so common in humans, and whether a selective advantage preserves this variation and the observed mito-nuclear interactions. As the mtDNA has high mutation rates with little or no recombination, it is prone to the accumulation of disabling mutations that could lead to its “meltdown” via Mueller’s ratchet (Lynch et al., 1993). However, mtDNA mutation followed by heteroplasmy is a requisite step in evolutionary adaptation. The identified nuclear QTLs for mtDNA heteroplasmy may represent mechanisms by which a reservoir of such variation can be tolerated and harnessed.

## Data Availability

In terms of data processed or generated as part of this study, we provide genetic association statistics for LD-independent lead SNPs and fine-mapped variants in UKB in addition to colocalization results (Supplementary tables 2-4). Full GWAS summary statistics from UKB and AoU will be made available in Zenodo upon peer-review. All GWAS sample sizes for each genetic ancestry group, meta-analysis, and phenotype can be found in Supplementary table 1. AoU policy does not currently permit public release of individual-level data due to important ethical and privacy considerations: https://www.researchallofus.org/wp-content/themes/research-hub-wordpress-theme/media/2020/05/AoU_Policy_Data_and_Statistics_Dissemination_508.pdf
In terms of external data used in this study, we leveraged GWAS summary statistics, and ancestry-specific LD-matrices, and a curated list of 29 common, high-quality disease phenotypes generated as part of the Pan UKBB project (Pan UKBB Initiative, 2022), with more information available online (https://pan.ukbb.broadinstitute.org). UKB phenotype and whole genome sequencing data can be accessed via the UKB Research Analysis Platform after completing a UKB access application: https://ukbiobank.dnanexus.com/landing. AoU phenotype and genotype data can be accessed via access to the Controlled Tier v6 on the AoU researcher workbench: workbench.researchallofus.org. Published mtscATACseq data used for chrM:302 analysis can be obtained via approval from dbGaP. Gene-sets for enrichment analyses can be obtained using COMPARTMENTS (https://compartments.jensenlab.org) and MitoCarta 2.0 (https://www.broadinstitute.org/files/shared/metabolism/mitocarta/human.mitocarta2.0.html) as described previously (Gupta et al., 2021). The GRCh37 and GRCh38 reference genomes as well as other standard reference data are available via the GATK resource bundle: https://gatk.broadinstitute.org/hc/en-us/articles/360035890811-Resource-bundle. Annotations for the baseline v1.1 and BaselineLD v2.2 models for S-LDSC as well certain other relevant reference data, including the HapMap3 SNP list, can be obtained from https://alkesgroup.broadinstitute.org/LDSCORE/. BLASTn was used as available from the NCBI: https://blast.ncbi.nlm.nih.gov/Blast.cgi. Known reference and polymorphic NUMTs were obtained from supplemental data as provided in published work (Calabrese et al., 2012; Dayama et al., 2014; Li et al., 2012; Wei et al., 2022).

## ACKNOWLEDGEMENTS

We thank Anna Kotrys, Tom Barton-Owen, and Carla Winter for thoughtful discussions, Melissa Walker for facilitating access to published data, Kristen Laricchia, Katherine Chao, Grace Tiao, Sinéad Chapman, and Namrata Gupta for help with gnomAD data and pipelines, and Wei Zhou for help with SAIGE. This project was supported in part by grants 5R35GM122455 (VKM) and 5F30AG074507 (RG) from the National Institutes of Health. VKM is an Investigator of the Howard Hughes Medical Institute. PFC is a Wellcome Trust Principal Research Fellow (212219/Z/18/Z), and a UK NIHR Senior Investigator, who receives support from the Medical Research Council Mitochondrial Biology Unit (MC_UU_00028/7), the Medical Research Council (MRC) International Centre for Genomic Medicine in Neuromuscular Disease (MR/S005021/1), the Leverhulme Trust (RPG-2018-408), an MRC research grant (MR/S035699/1), an Alzheimer’s Society Project Grant (AS-PG-18b-022). This research was supported by the NIHR Cambridge Biomedical Research Centre (BRC-1215-20014). The views expressed are those of the author(s) and not necessarily those of the NIHR or the Department of Health and Social Care. This research has been conducted using the UK Biobank Resource under Application Number 31063. The All of Us Research Program is supported by the National Institutes of Health, Office of the Director: Regional Medical Centers: 1 OT2 OD026549; 1 OT2 OD026554; 1 OT2 OD026557; 1 OT2 OD026556; 1 OT2 OD026550; 1 OT2 OD 026552; 1 OT2 OD026553; 1 OT2 OD026548; 1 OT2 OD026551; 1 OT2 OD026555; IAA #: AOD 16037; Federally Qualified Health Centers: HHSN 263201600085U; Data and Research Center: 5 U2C OD023196; Biobank: 1 U24 OD023121; The Participant Center: U24 OD023176; Participant Technology Systems Center: 1 U24 OD023163; Communications and Engagement: 3 OT2 OD023205; 3 OT2 OD023206; and Community Partners: 1 OT2 OD025277; 3 OT2 OD025315; 1 OT2 OD025337; 1 OT2 OD025276. In addition, the All of Us Research Program would not be possible without the partnership of its participants.

## DATA AVAILABILITY

In terms of data processed or generated as part of this study, we provide genetic association statistics for LD-independent lead SNPs and fine-mapped variants in UKB in addition to colocalization results (**Supplementary tables 2-4**). Full GWAS summary statistics from UKB and AoU will be made available in Zenodo upon peer-review. All GWAS sample sizes for each genetic ancestry group, meta-analysis, and phenotype can be found in **Supplementary table 1**. AoU policy does not currently permit public release of individual-level data due to important ethical and privacy considerations: https://www.researchallofus.org/wp-content/themes/research-hub-wordpress-theme/media/2020/05/AoU_Policy_Data_and_Statistics_Dissemination_508.pdf In terms of external data used in this study, we leveraged GWAS summary statistics, and ancestry-specific LD-matrices, and a curated list of 29 common, high-quality disease phenotypes generated as part of the Pan UKBB project (*Pan UKBB Initiative*, 2022), with more information available online (https://pan.ukbb.broadinstitute.org). UKB phenotype and whole genome sequencing data can be accessed via the UKB Research Analysis Platform after completing a UKB access application: https://ukbiobank.dnanexus.com/landing. AoU phenotype and genotype data can be accessed via access to the Controlled Tier v6 on the AoU researcher workbench: workbench.researchallofus.org. Published mtscATACseq data used for chrM:302 analysis can be obtained via approval from dbGaP. Gene-sets for enrichment analyses can be obtained using COMPARTMENTS (https://compartments.jensenlab.org) and MitoCarta 2.0 (https://www.broadinstitute.org/files/shared/metabolism/mitocarta/human.mitocarta2.0.html) as described previously (Gupta et al., 2021). The GRCh37 and GRCh38 reference genomes as well as other standard reference data are available via the GATK resource bundle: https://gatk.broadinstitute.org/hc/en-us/articles/360035890811-Resource-bundle. Annotations for the baseline v1.1 and BaselineLD v2.2 models for S-LDSC as well certain other relevant reference data, including the HapMap3 SNP list, can be obtained from https://alkesgroup.broadinstitute.org/LDSCORE/. BLASTn was used as available from the NCBI: https://blast.ncbi.nlm.nih.gov/Blast.cgi. Known reference and polymorphic NUMTs were obtained from supplemental data as provided in published work (Calabrese et al., 2012; Dayama et al., 2014; Li et al., 2012; Wei et al., 2022).

## CODE AVAILABILITY

We release the full WDL pipelines for mtDNA analysis from whole genome sequencing data on GitHub: (https://github.com/rahulg603/mtSwirl). We also provide the code we used to run the pipeline on the UKB Research Analysis Platform, AoU, and Terra, consolidate all data, and perform mtDNA sample and variant QC. See **Methods** and the README for more information on how to use the pipeline. Several tools were used as part of mtSwirl, including GATK v4.2.6.0 (https://gatk.broadinstitute.org/), samtools v1.9 (https://github.com/samtools/samtools) and bcftools v1.16 (https://github.com/samtools/bcftools), Haplochecker 0124 (https://github.com/genepi/haplocheck), R (r-project.org), Hail (hail.is), and UCSC kent LiftOver tools (genome-source.soe.ucsc.edu/kent.git).

We used several published tools and scripts to perform downstream analysis of the mtDNA callset in this study. All data wrangling, statistical analysis, and figure generation was performed using either Hail v0.2.98 (hail.is) or R v4.2.1 (r-project.org). Parallelization of tasks in UKB was performed using Hail Batch (batch.hail.is) and in AoU using Cromwell v77 (cromwell.readthedocs.io). GWAS was performed in UKB using SAIGE v1.1.5 (saigegit.github.io). For scaling of UKB GWAS, a custom modification of the GWAS pipeline from the Pan UKBB pan-ancestry GWAS was used (https://github.com/atgu/ukbb_pan_ancestry). GWAS was performed in AoU using Hail. mtDNA PCA was performed in R using the irlba v2.3.5.1 package (https://cran.r-project.org/web/packages/irlba/index.html). Multinomial models were trained using the nnet v7.3-17 package in R (https://cran.r-project.org/web/packages/nnet/index.html). Circos plots were made using the circlize package v0.4.15 in R (https://jokergoo.github.io/circlize_book/book/). For analysis of chrM:302 in single cell data, we used BedTools v2.29.2 (bedtools.readthedocs.io). LD clumping was performed using Plink v1.90 (https://www.cog-genomics.org/plink/). Finemapping was performed using FINEMAP-inf and SuSiE-inf (https://github.com/FinucaneLab/fine-mapping-inf). eQTL data was obtained from GTEx v8 (gtexportal.org) and the eQTL catalogue release 4 (https://www.ebi.ac.uk/eqtl/). For replication analysis effect size comparisons, the deming pacakge v1.4 was used in R (https://cran.r-project.org/web/packages/deming/index.html). Heritability estimates and enrichment analyses were performed using stratified LD-score regression (https://github.com/bulik/ldsc).

## METHODS

### Overview of mtSwirl

Here we develop mtSwirl, a scalable pipeline for mitochondrial DNA copy number and variant calling which makes calls relative to an internally generated per-sample consensus sequence before mapping all calls back to GRCh38. In addition to GRCh38 reference files and whole-genome sequencing (WGS) data, the mtSwirl pipeline takes as input nuclear genome reference intervals that represent regions with high homology to the mtDNA (reference NUMTs). We constructed a set of 385 putative NUMTs by using a BLAST-based inventory of reference NUMTs published previously (Li et al., 2012), extending the boundaries of each interval by 500 bases, and merging any overlapping intervals. Initial variant calls within the mtDNA and reference NUMT regions are made from mapped WGS data using Mutect2 and HaplotypeCaller respectively (via GATK v4.2.6.0), and haplogroup inference is performed via Haplogrep (Weissensteiner et al., 2016). Consensus sequences are subsequently constructed using homoplasmies (mtDNA) and homozygous alternate (nucDNA) calls. Reads are realigned to the new consensus sequence and variants are called on the mtDNA using Mutect2. To avoid the artificial coverage depression at the ends of the mtDNA reference genome, we call variants in the control region after alignment to a shifted mtDNA molecule. All variant calls and per-base coverage estimates are then returned to GRCh38 coordinates and output from the pipeline. See **Supplementary note 1** for more details. We release two versions of our pipeline on GitHub (https://github.com/rahulg603/mtSwirl): mtSwirlSingle, a single-sample pipeline intended for use with Cromwell and on platforms with high worker limits like Terra and the AllofUs Workbench, and mtSwirlMulti, a multi-sample version which processes multiple samples serially per machine intended for use on platforms with a smaller parallel worker limit such as the UKB Research Analysis Platform (RAP).

### Cohorts

#### UK Biobank (UKB)

The UK Biobank is a large prospective cohort study of ∼500,000 individuals in the UK (Sudlow et al., 2015), ∼200,000 of whom had whole genome sequencing performed at the time of this study. Samples were selected for the first round of WGS using a pseudorandom approach to ensure that included samples were representative of the full cohort. Sequencing data was generated using DNA extracted from buffy coat obtained from participants; more details have been reported previously (Halldorsson et al., 2022). All UKB data was accessed under application 31063 and mtDNA variant calling was performed on the UKB RAP.

#### AllofUs (AoU)

AllofUs is a large longitudinal cohort study based in the United States, with a central goal of enrolling a diverse cohort of participants providing electronic health record data over time, specimens for genetic analysis, survey responses, and standardized biometric measurements (“The ‘All of Us’ Research Program,” 2019). At the time of this study, 98,590 individuals had completed whole genome sequencing on samples obtained from whole blood. DNA extraction was completed at the Mayo Clinic, and sequencing was performed at three sequencing centers (Baylor College of Medicine, Broad Institute, and University of Washington) using harmonized protocols. Post-sequencing variant and sample quality control was performed by the AllofUs Data and Research Center (DRC). All mtDNA analyses were performed using the AllofUs Researcher Workbench in the Controlled Tier v6 workspace: “Genetic determinants of mitochondrial DNA phenotypes” using data from the Q2 2022 release. See https://support.researchallofus.org/hc/en-us/article_attachments/7237425684244/All_Of_Us_Q2_2022_Release_Genomic_Quality_Report.pdf for more details on genomics QC and pre-processing.

#### gnomAD v3.1 subset

gnomAD v3.1 is a database aggregating whole genome sequencing data from 76,156 samples from several experiments and projects around the world, as part of which an mtDNA variant callset was recently produced (Laricchia et al., 2022). Samples were sourced from several study designs including case-control studies for common diseases, population-based cohorts, and observational studies. Individuals with inborn severe pediatric disease were excluded. Most data are sourced from sequencing performed on either blood samples extracted using study-specific methodologies or from cell lines (Laricchia et al., 2022). We made use of a subset of the gnomAD v3.1 samples to prototype our pipeline (mtSwirl) and compare its performance to previous mtDNA copy number and variant calls (“Vanilla”). We excluded samples with very high mtDNA copy number as done previously (Laricchia et al., 2022) as these are likely cell line samples and not from whole blood; we used a more stringent threshold of 350 as we wanted to maximally enrich for whole blood samples for this trial. We also removed samples with mtCN < 50 due to elevated NUMT contamination in these samples (Laricchia et al., 2022, **Supplementary figure 7C**). We selected ∼6300 samples from gnomAD v3.1 to maximize inclusion of diverse haplogroups including those underrepresented in UK Biobank (**Supplementary figure 2A**). We specifically supplemented samples belonging to the L haplogroups and enforced a cap on the number of samples assigned to either NFE (Non-Finnish European) or FIN (Finnish). For other larger haplogroups we performed random subsampling proportional to the original composition of the gnomAD dataset to achieve our final sample size. All analyses were performed using Terra (*Terra*, n.d.).

### Computing mean nuclear DNA coverage in UKB

As mean nuclear DNA coverage was not available in UK Biobank, we used samtools v1.9 idxstats (Danecek et al., 2021), samtools flagstat, and GATK v4.2.6.0 CollectQualityYieldMetrics as part of the mtSwirlMulti pipeline to efficiently and economically estimate mean coverage on the nuclear DNA. idxstats-based counts of total mapped reads were computed over autosomes with the subsequent formula applied to get average nuclear DNA coverage after removing contributions from duplicate reads:

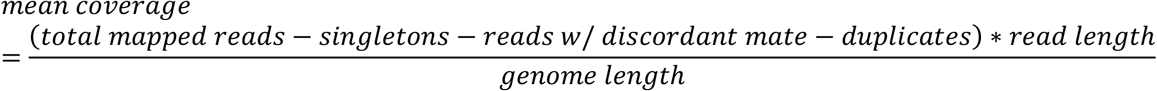

### Computing mtDNA copy number

Across all cohorts we use the following formula to compute mtDNA copy number:

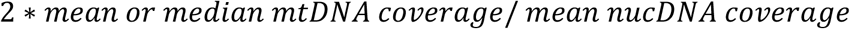

We default to use of mean mtDNA coverage for main mtCN-related analyses.

### Post-calling mtDNA phenotype QC

To integrate our variant calls and perform sample and variant QC, we extended a previously developed pipeline (Laricchia et al., 2022). Single-sample VCFs emitted from mtSwirl were merged into a single Hail MatrixTable (v0.2.98; (Hail Team, n.d.)) upon which all downstream steps were conducted.

For sample QC, any samples showing homoplasmic variant overlap (see **Supplementary note 1**) were removed. We observed a significant elevation in heteroplasmic SNV calls among samples with mtCN below 50, with a stabilization of heteroplasmic calls above 50 mtDNA copies per cell (**Supplementary figure 7C**), highly suggestive of elevated NUMT contamination in the low copy number samples. Thus, to avoid contamination of our results, all samples with mtCN < 50 were removed. Finally, all samples with evidence of contamination > 2% were removed, as estimated by either (1) mtDNA contamination via Haplocheck 0124 (Weissensteiner et al., 2021) in mtSwirl, (2) nucDNA contamination, or (3) the presence of multiple haplogroup-defining variants at abnormally low allele fraction. Given the small count of samples processed in 2006 and abnormally elevated mtDNA copy number estimates in these samples (**Supplementary figure 3E**), we excluded these samples from all UKB analyses.

For variant QC, (1) variants with a very low heteroplasmy (< 0.01) were called as reference with a heteroplasmy of 0, (2) variants with heteroplasmy below 0.05 were flagged and removed as these are at high risk of being enriched for NUMT-derived signals, and (3) all variant calls flagged by Mutect2 were removed. For all sites, a minimum coverage threshold of 100 was used to distinguish between homoplasmic reference calls and sites without variant calls due to low variant-calling confidence as done previously (Laricchia et al., 2022). mtDNA variants were annotated using the Variant Effect Predictor (VEP) v101 (McLaren et al., 2016) and dbSNP v151 (Sherry et al., 1999). Variants with at least 0.1% of samples passing filters showing a heteroplasmy between 0 and 0.5 were annotated as “common low heteroplasmy”. Variant calls failing QC were coded with a missing heteroplasmy.

For mtCN, we remove the samples identified during variant callset sample QC showing signs of contamination, abnormal overlapping homoplasmy calls, or which were processed in 2006. Since we expect mtDNA-wide coverage measures, such as mtCN, to be robust to NUMTs, we do not enforce hard cutoffs on mtCN measurements.

### Construction of mtDNA heteroplasmy phenotypes

We defined our set of common heteroplasmies in UKB as “common low heteroplasmy” variants (**Methods**) which are present as heteroplasmies in at least 500 individuals, resulting in 39 variants. We produced two main sets of phenotypes: (1) a “case-only” dataset consisting of heteroplasmy values for these variants where any individuals without the variant detected were coded as missing and (2) as “case-control” dataset where cases consisted of those with any detectable heteroplasmy and controls consisted of those with the variant not detected. In both phenotype schemes, samples identified as homoplasmic for each variant were always coded as missing. For the case-control dataset, only samples which could be accurately inferred as reference for each variant were labeled as controls – specifically, the sample was coded as missing for a variant if it had a coverage < 100 at the site or showed the variant call as QC-fail (**Methods**).

For sensitivity analyses, we produced several additional case-only heteroplasmy datasets: (1) where any variant calls supported by an alternate allele depth (AD alt) of less than the mean nuclear DNA coverage of the sample were made missing; (2) where heteroplasmy estimates were corrected for the depth of mtDNA coverage at the variant site after realignment; and (3) where length heteroplasmy estimates at chrM:302 were corrected for median coverage at CSBII. All corrections were performed by obtaining residuals from the linear regression of the heteroplasmy onto the covariate for each variant across all samples prior to genetic analysis.

### mtDNA phenotype covariate correction approach

We investigated time of day of blood draw, fasting time, assessment date, and assessment center as technical covariates for mtDNA traits. As draw time and assessment date are continuous, we used natural splines in the correction model to flexibly model nonlinear relationships between these covariates and the mtDNA phenotype. We used knots placed roughly seasonally to model seasonal variation in mtDNA phenotypes – these corresponded to 3-month increments starting on July 1^st^ 2007 and ending on July 1^st^ 2010. For draw time, we used a natural spline basis with 5 degrees of freedom. Assessment month and assessment center were modeled as indicator variables. Fasting times were provided in increments of 1 hour and thus were modeled as indicator variables; fasting times of > 18 hours were labeled as 18 and fasting times of 0 were labeled as 1. All terms were included in a joint model for correction.

We also investigated the relationship between mtDNA phenotypes and blood cell type percentages and mean blood cell volumes. We selected all non-redundant traits available: white blood cell leukocyte count, haematocrit percentage, platelet crit, monocyte percentage, neutrophil percentage, eosinophil percentage, basophil percentage, reticulocyte percentage, high light scatter reticulocyte percentage, immature reticulocyte fraction, mean corpuscular volume, mean reticulocyte volume, mean sphered cell volume, mean platelet thrombocyte volume. We did not include nucleated red blood cell percentage as only ∼1% of the entire UKB cohort has non-zero values for this measure, and we excluded lymphocyte percentage given collinearity with neutrophil percentage (r = 0.92) and the sum-to-1 property of the white blood cell (WBC) differential measurements. To avoid excess leverage from outlying blood cell measurements, we removed any blood measurements with a Z-score > 4. All terms were included in a joint model for correction.

For both the technical covariate and blood cell type models, F-test p-values were obtained for each of the 40 mtDNA phenotypes (39 case-only heteroplasmies and mtCN). For any phenotypes which showed F-test p-values < 0.05/40 (Bonferroni corrected), we produced corrected versions of the phenotype by obtaining the residuals from the regression of the mtDNA phenotype onto covariates of interest prior to genetic analysis. For mtDNA copy number, adjustments were performed with log(mtCN) as the response variable. For heteroplasmy estimates, adjustments were performed with case-only heteroplasmies as the response variable. The specific corrections implemented were:

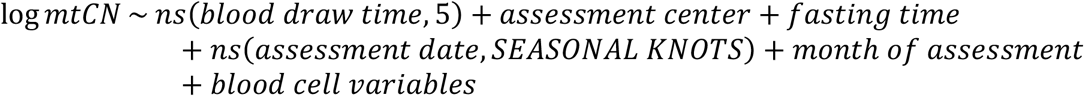

As sensitivity analyses for case-only heteroplasmy phenotypes, residuals from the following models were produced:

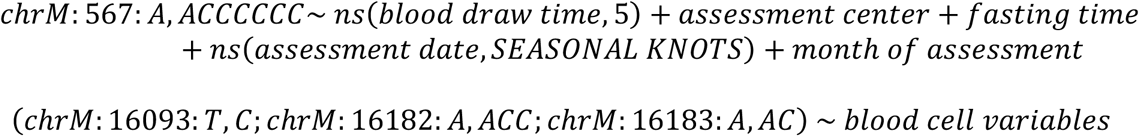

For each response variable, residuals were generated using `residuals(lm(model))` as implemented in R v4.2.1. In all visualizations of corrected variables (e.g., mtCN_corr_), we rescale the residualized variable by adding the pre-corrected mean. In the case of mtCN_corr_, we rescale the residualized variable and then exponentiate the same to return corrected values back to an absolute scale. See **Supplementary note 2** and **3** for more details.

### mtDNA PCA and predictive power for mtDNA haplogroups

To construct a high-quality variant genotype matrix for PCA, we obtained the set of homoplasmic variants (heteroplasmy >= 0.95) passing QC identified at a MAF >= 0.001 in UKB. Any samples with a QC-pass homoplasmy detected were coded as 1 for each respective variant; all others were coded as 0. This binary genotype matrix was subsequently filtered to the set of unrelated samples upon which we computed the first 50 principal components after centering and scaling using the efficient truncated singular value decomposition algorithm implemented in the `irlba` v2.3.5.1 package in R. Related samples were projected onto these PCs to produce a set of mtDNA-PC coordinates for each sample. The set of related samples were defined previously in the Pan UKBB project (*Pan UKBB Initiative*, 2022). In brief, PC-relate was used as implemented in Hail within each assigned genetic ancestry group in UKB and the maximal set of unrelated samples were identified via the maximal independent set algorithm implemented in Hail.

To assess the goodness of fit of mtDNA PCs for the prediction of top-level mtDNA haplogroups, we fit a multinomial model with top-level haplogroup as the response variable and the first 30 mtDNA PCs as explanatory variables as implemented in the `nnet` v7.3-17 package in R (Venables & Ripley, 2002). We only included samples belonging to haplogroups with at least 30 samples in UKB. For assessment of the predictive power of mtDNA PCs for “level 2” haplogroups, we fit multinomial models using a similar approach within each top-level haplogroup, with “level 2” haplogroups as the response variable. In all cases, a null model was fit in parallel with the same response variable with only an intercept term. We computed McFadden’s pseudo-R^2^ for each model via the following formula:

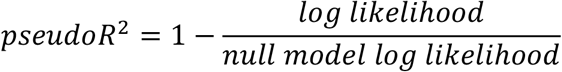

### Correlations between mtCN, mtCN_corr_, blood cell composition, heteroplasmies, and disease phenotypes

We obtained common disease diagnoses from UKB via a previously curated set of phecodes and ICD10 codes corresponding to major common diseases (*Pan UKBB Initiative*, 2022) along with demographic variables (age, sex) and blood cell composition phenotypes (**Methods**). We obtained mtCN_raw_, mtCN_corr_, common (N > 500) case-only heteroplasmies (**Methods**), and three major blood cell composition traits (platelet crit, monocyte count, and neutrophil count) and performed z-score transformation for each. To test for associations with disease phenotypes, we used a logistic regression model via the `glm` function in R, including age, sex, age_2_, age_2_*sex, age*sex, top-level haplogroup, and genetic ancestry group assignment as covariates:

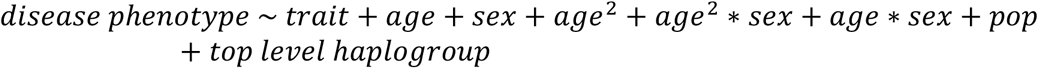

We included haplogroups with at least 30 individuals represented in UKB. Odds ratios were obtained as exp (*β*_*trait*_), and the 95% CI was obtained as exp (*β*_*trait*_ ± 1.96 ∗ *SE*_*trait*_).

### Derivation of mtDNA coverage discrepancy phenotypes

We obtained mtDNA intervals corresponding to the 7s DNA, heavy strand origin, CSBII, CSBIII, and the light strand promoter (LSP) (Falah et al., 2017; Tan et al., 2016; Xuan et al., 2006). We computed per-individual median mtDNA coverages within the regions corresponding to the first third of the 7s DNA (“7s DNA”), the region between CSBII and the heavy strand origin (“7s DNA primer”), and the region between CSB III and the LSP (“7s RNA primer”). To generate coverage discrepancy phenotypes, we regressed DNA primer coverage onto either 7s DNA coverage or 7s RNA primer coverage. To avoid coverage discrepancies attributable to inherited mtDNA variation within the regions of interest, we included indicator variables for all top-level haplogroups with at least 30 samples as well as their interactions with 7s DNA or 7s RNA primer coverage. The residuals from the following model were used as the coverage discrepancy phenotype for GWAS:

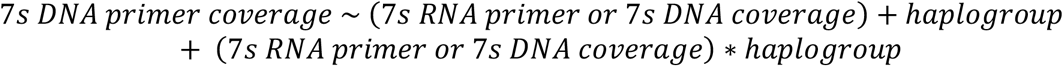

### Relatedness analyses in UKB

Relatedness was computed and sibling-sibling and parent-offspring pairs were inferred as previously described in UKB (Karczewski et al., 2022). For the assessment of transmission of all QC-pass mtDNA variants, we restricted to only variants found in 5 or more samples.

### Determination of chrM:302 length heteroplasmy composition

To construct length heteroplasmy profiles, we obtained all post-QC variant calls made at position chrM:302. We defined a “reference” call at chrM:302 for each sample as 1 − *sum(heteroplasmy of any allele at chrM*: 302). All samples without variant calls at chrM:302 were assigned a reference fraction of 1, with samples with a depth of < 100 at chrM:302 (after local re-alignment during variant calling) excluded. For each sample, we combined all heteroplasmies from calls other than reference, chrM:302:A,AC, chrM:302:A,ACC, and chrM:302:A,ACCC into an “Other” category. Any calls with a missing value for a chrM:302 allele were imputed as a heteroplasmy of 0 for the purposes of visualizations and analyses.

### Associations between pathogenic variant carrier status and continuous phenotypes in UKB

We obtained continuous phenotypes available in UKB corresponding to classic symptoms of MELAS – diabetes-like symptoms (elevated triglycerides (ID 30870), elevated hemoglobin A1c (ID 30750)) and hearing impairment (via the speech-reception-threshold estimate (IDs 20019 and 20021)) – as well as the results from the visual acuity test for analysis of known pathogenic variants for Leber’s hereditary optic neuropathy (logMAR from visual acuity test (IDs 5201 and 5208)). All obtained phenotypes were filtered to samples with available mtDNA variant calls and corrections were applied for age, sex, age_2_, age_2_*sex, age*sex, and genetic ancestry group assignment by obtaining residuals from the following linear regression model using `residuals(lm(model))` in R:

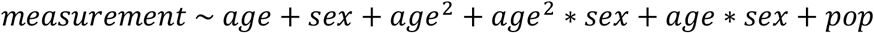

As blood biomarkers tend to have log-normal distributions, corrections were applied after log transformation of HbA1c and triglyceride levels. Post-correction, all measurements were returned to their original scale by adding the pre-correction dataset-wide means for each measurement modality. Final estimates for the speech-recognition-threshold and vision logMAR were generated by averaging measurements for the left and right ear and eye respectively.

Carriers of known pathogenic mtDNA variants were defined as individuals carrying the variant post-QC at any fraction. We defined a set of controls as individuals with none of the ten known pathogenic mtDNA variants tested. Only samples which could be accurately inferred as reference for all ten variants were labeled as controls – the sample was excluded if, for any of the ten variants, it had a coverage of below 100 at the site or showed a QC-fail variant call (**Methods**).

Comparisons between residual phenotype values among variant carriers versus global controls were performed only for variant-phenotype pairs with more than 10 defined phenotype values among variant carriers. P-values were obtained by performing a two-sample t-test between phenotype values among variant carriers and the set of global controls, and q-values were obtained by applying the Benjamini-Hochberg procedure.

### Creation of mutational spectrum categories

Heteroplasmic SNV mutation types in AllofUs were constructed using the set of QC-pass heteroplasmic SNVs. For each SNV type, the set of individuals without any heteroplasmic variants was identified as those with no QC-pass variant call of that type; these individuals were included as zeros in estimates of the mean SNV count of each type.

### chrM:302 length heteroplasmy inference in single cells

We used the BedTools (Quinlan & Hall, 2010) intersect tool (v2.29.2) to identify read alignments completely spanning the chrM:300-318 locus in the mtscATAC-seq data from Walker et al., 2020, obtained with Massachusetts General Hospital IRB approval under protocol #2016P001517. We then iterated over these reads and classified their chrM:302 length variant by extracting the poly-C/G tracts using a regular expression, ‘AA(CCC+[CT]CC+)GC’, anchored on the two constant bp on either side of the variant region to detect the canonical variant structure of two poly-C/G tracts with or without a single intervening A/T. Alleles in matching reads were classified based on the length of their poly-C/G tracts, while alleles in the reads that did not match the regular expression were classified as NA. Next, we filtered out any reads with cell barcodes that were not in the published list of cell calls, and further restricted our analysis to only the cells with at least 20 reads at the chrM:300-318 locus. For each of these high coverage cells, we calculated the fraction of reads showing each of the top three most common length variants (G_6_AG_8_, G_6_AG_9_, and G_6_AG_10_) and aggregated any other detected alleles into the remainder (other) for display as a stacked bar plot. We also estimated bulk heteroplasmy by summing the allele counts from the high coverage cells and re-calculating the fractions for the top three length variants, again with all other alleles being aggregated into the remainder “other” category.

### UKB GWAS approach

All GWAS was performed in UKB using approaches as performed in the Pan UKBB initiative (*Pan UKBB Initiative*, 2022). In brief, ancestry assignment was performed by projecting UKB samples into genotype PC-space constructed from reference samples from 1000 Genomes (1KG) phase 3 and the Human Genome Diversity Project (HGDP) and subsequently using a random forest classifier to assign continental labels trained on the 1KG+HGDP reference data. Within each ancestry group, PCA was performed among unrelated samples with related samples projected onto this PC-space. Further sample QC was performed excluding samples as described as part of the Pan UKBB initiative (*Pan UKBB Initiative*, 2022), including removal of ancestry outliers using a centroid-based metric, individuals with high genotype missingness, sex discordance, and sex chromosome aneuploidies. Variant QC was also performed on UKB-provided imputed v3 variants as part of the Pan UKBB initiative (*Pan UKBB Initiative*, 2022), including only those with INFO scores > 0.8 on autosomes and the X-chromosome. Association tests were performed only on variants with a minor allele count (MAC) > 20.

For GWAS, SAIGE v1.1.5 (Zhou et al., 2018) was used to perform association tests within each assigned ancestry group using the first 10 per-population PCs, age, age*sex, age^2^, and age^2^*sex as covariates (referred to as “baseline”). Ancestry groups were only included if at least 50 individuals had the phenotype defined. The use of the SAIGE GRM-based approach allowed for the inclusion of related samples in the GWAS, and we enabled leave-one-chromosome-out fitting in all steps. For all continuous phenotype GWAS (case-only mtDNA heteroplasmy traits and mtCN), phenotypes were inverse rank normalized prior to genetic analysis.

For all main mtDNA heteroplasmy analyses, top-level mtDNA haplogroup was included as an additional set of covariates in the GWAS model as a set of 24 indicator variables with haplogroup A as reference. Any samples belonging to top-level haplogroups with fewer than 30 samples represented were excluded. The same GWAS model was used for sensitivity analysis of case-only heteroplasmies after removing calls with AD alt < mean nucDNA coverage, after correction for local variant coverage, after correction for CSBII coverage, and after correction for technical or blood trait covariates (**Methods**). For the main mtCN analyses, we used only the baseline covariates to perform genetic associations with mtCN_raw_ and mtCN_corr_.

We performed two additional sensitivity analyses for case-only heteroplasmy GWAS: (1) inclusion of 30 mtDNA PCs as covariates in the GWAS model instead of top-level haplogroup for 7 variants which showed relatively high heterogeneity across level 2 haplogroups, and (2) inclusion of mtCN as a covariate in the GWAS model for all case-only heteroplasmies in addition to top-level haplogroup. We also tested the effects of including top-level haplogroup indicator variables as additional covariates in GWAS for mtCN_raw_ and mtCN_corr_.

### AllofUs GWAS approach

We performed GWAS in AllofUs as replication for our main case-only heteroplasmy analyses in UKB. Ancestry inference was performed upstream by the AllofUs Data and Research Center (DRC). In brief, AoU samples were projected into the PCA space of genotypes from chromosomes 20 and 21 from HGDP and 1KG, and a random forest classifier trained to identify ancestry labels in 1KG+HGDP was used to assign AoU samples continental ancestry labels.

We performed sample and variant QC after WGS variant calls were imported into Hail. Multi-allelic sites were split and sites with very low pre-computed AF were removed (MAF > 0.0001 retained). For sample QC, samples flagged by the DRC as population outliers for several metrics or identified as related by the DRC were excluded. For variant QC, we removed any variants filtered by the DRC, which occurred in brief because of no high-quality genotypes for the variant (defined as GQ >= 20, DP >= 10, AB >= 0.2 for heterozygotes), excess heterozygotes, or a low quality score for the variant. We further removed any variants not in Hardy-Weinberg equilibrium (one-sided p <= 1e-10) and variants with a call rate <= 0.95. Finally, we removed any variants with MAC < 20 in each assigned ancestry group.

We next extracted covariates relevant for our GWAS model. We used a SQL query to obtain date of birth in the controlled data repository and used the provided QC flat files to obtain sex assigned at birth. As date of sample collection was not provided, approximate age was constructed for all analyses by subtracting the year of birth from the year 2021. To address residual stratification within assigned ancestry groups, we produced PCs within each ancestry group using a very similar approach as used in UKB (**Methods**) as we found that the provided PCs did not appropriately handle stratification among positive control phenotypes like height, blood glucose, diastolic blood pressure, and systolic blood pressure (**Supplementary note 4**). We included 20 recomputed PCs, in addition to approximate age, age^2^, age*sex, and age^2^*sex as covariates in the final GWAS model. We did not perform genetic analysis for the MID group as less than 400 samples with available WGS data were assigned MID.

We used Hail with the `hl.linear_regression_rows()` method to perform GWAS after all QC. As described in **Methods**, we performed genetic analysis for all QC-pass case-only mtDNA heteroplasmies with homoplasmic calls set to missing. As this analysis is intended for replication, we included any variants found in 300 or more samples across any ancestry group, resulting in 41 variants for genetic analysis. Of these, 36 were also analyzed in UKB; 3 UKB variants were not sufficiently common in AoU for genetic analysis. As in UKB, for the analysis of case-only mtDNA heteroplasmies, top-level mtDNA haplogroup was included as covariates in the GWAS model as a set of 27 indicator variables in addition to age, sex, and PC covariates. Samples belonging to top-level haplogroups with fewer than 30 samples in AoU were excluded. All case-only mtDNA heteroplasmy phenotypes were inverse rank normalized prior to analysis.

See the AllofUs genotype quality report for more information on upstream genotype data and sample QC, ancestry inference, and relatedness inference (https://support.researchallofus.org/hc/en-us/article_attachments/7237425684244/All_Of_Us_Q2_2022_Release_Genomic_Quality_Report.pdf).

### Heritability estimation and enrichment analyses for mtCN

Stratified linkage disequilibrium score regression (S-LDSC, Finucane et al., 2015) was used for heritability estimation and enrichment analyses for mtDNA copy number in UKB as performed previously (Gupta et al., 2021). In brief, we analyzed EUR summary statistics in UKB, restricting variants to those in HapMap3 (HM3). We estimated overall SNP-heritability controlling for 97 annotations corresponding to coding regions, enhancer regions, minor allele frequency bins, and others (Gazal et al., 2017, referred to as baselineLD v2.2). For enrichment analyses, we obtained gene-sets corresponding to (1) the top 10% of genes specifically expressed in major tissues from GTEx (Finucane et al., 2018) and (2) genes producing protein products that localize to each major organelle with high confidence using COMPARTMENTS (Binder et al., 2014). Variants were mapped to each gene with a 100kb symmetric window and LD scores for each gene-set annotation were computed using the 1000G EUR reference panel (https://alkesgroup.broadinstitute.org/LDSCORE/). Heritability enrichment for all gene-sets was tested using S-LDSC atop the baseline v1.1 model, controlling for 53 annotations including coding regions and 5’ and 3’ UTRs (Finucane et al., 2015).

### Cross-ancestry meta-analysis in UKB and AllofUs

We conducted a fixed-effect meta-analysis across ancestries in each cohort (UKB and AoU) based on inverse-variance weighted betas and standard errors (de Bakker et al., 2008). For each ancestry, we excluded low-confidence variants defined as MAC <= 20 in either biobank. We computed effect size heterogeneity P-values across ancestries using Cochran’s Q-test (Cochran, 1954). All computation was done using Hail v0.2.

All visualizations of main GWAS (e.g., mtCN, coverage discrepancy traits, heteroplasmy traits) are of cross-ancestry meta-analyses after restriction to the set of “high quality” variants as defined previously (*Pan UKBB Initiative*, 2022).

### Identification of LD-independent lead SNPs and locus definitions

Clumping was performed using Plink v1.90 (Purcell et al., 2007) in Hail Batch for GWAS results obtained in UK Biobank after filtering to high quality variants. We used significance thresholds of 1 for both the index and clumped SNPs, set the LD threshold for clumping at 0.1, and set the distance threshold at 500kb. We used single ancestry and multi-ancestry LD reference panels corresponding to the ancestry groups included in the final multi-ancestry meta-analyses for each mtDNA phenotype as well as for blood cell traits. Reference panels were constructed by randomly sampling 5000 individuals from all samples within any given set of ancestry groups in the UK Biobank. For the single-ancestry LD panels corresponding to ancestry groups with less than 5000 individuals (EAS and MID), the full sample available for each ancestry group was used. More details on the LD reference panels can be found as part of the Pan UKBB project (*Pan UKBB Initiative*, 2022). Clumping output files from Plink were converted to Hail Tables and then combined into MatrixTables using the multi-way-zip-join method as implemented in Hail. We defined distinct loci conservatively by starting with genome-wide significant LD-independent lead SNPs and merging any SNPs within 2 Mb of one another.

### Replication of previous mtCN GWAS with our study

We performed a comparison of significant loci identified in a previous GWAS of mtCN in UKB (Longchamps et al., 2021) with our own by performing LD clumping on previously released summary statistics as described (**Methods**) using 1KG phase 3 EUR reference data for LD. We defined distinct loci as described (**Methods**), merging any SNPs within 2 Mb of one another, arriving at 96 loci previously identified. We defined a replicated locus with mtCN_raw_ or mtCN_corr_ as one where our GWAS showed a signal at p < 5*10^−5^ or 5*10^−8^ within 2 Mb of the most significant variant identified in the previous study within each locus.

### Bidirectional Mendelian randomization between UKB mtCN and neutrophil count

GWAS effect sizes and LD-independent loci from the UKB cross-ancestry meta-analysis for raw mtCN and fully corrected mtCN were obtained. Summary statistics and LD-independent loci from GWAS among EUR for neutrophil count (ID 30140) were obtained from the Pan UKBB project (*Pan UKBB Initiative*, 2022). Sites for comparison were restricted to those passing variant QC as performed in UKB (**Methods**). For each mtCN phenotype, neutrophil count and mtCN GWAS effect sizes were obtained for all mtCN genome-wide significant variants, and vice-versa, mtCN and neutrophil count GWAS effect sizes were obtained for all neutrophil count genome-wide significant variants. We assessed the relationship between pre- and post-correction mtCN GWAS effect sizes and neutrophil count GWAS effect sizes via inverse-variance weighted linear regression using weights corresponding to 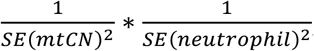, where effect size standard errors were obtained from the respective GWAS.

### Fine-mapping in UKB

To identify putative causal variants in associated loci, we conducted statistical fine-mapping of mtDNA traits in UKB using cross-ancestry meta-analysis summary statistics. While we previously showed that fine-mapping a meta-analysis is often miscalibrated due to heterogeneous characteristics of constituent cohorts (e.g., genotyping or imputation) (Kanai et al., 2022), a within-cohort cross-ancestry meta-analysis like the present study is a notable exception given no such heterogeneity systematically exists across ancestries.

We used FINEMAP-inf and SuSiE-inf which model infinitesimal effects (Cui et al., 2022), with cross-ancestry meta-analysis summary statistics (**Methods**) and a covariate-adjusted in-sample dosage LD matrix (Kanai et al., 2021). We defined fine-mapping regions based on a 3 Mb window around each lead variant and merged regions if they overlapped as described previously (Kanai et al., 2021). We excluded the major histocompatibility complex (MHC) region (chr 6: 25–36 Mb) from analysis due to extensive LD structure in the region. For each method, we allowed up to 10 causal variants per region and derived posterior inclusion probabilities (PIP) of each variant using a uniform prior probability of causality. To achieve better calibration, we computed min(PIP) across the methods and derived up to 10 independent 95% credible sets (CS) from SuSiE-inf as described elsewhere (Kanai et al., 2021). All reported PIP are min(PIP) between the two methods.

### Enrichment of functional categories among fine-mapped variants

We computed functional enrichment of fine-mapped variants across the mtDNA traits in UKB. We first annotated each variant with seven functional categories (pLoF, missense, synonymous, 5’ UTR, 3’ UTR, promoter, cis-regulatory element [CRE], and non-genic) as described previously (Kanai et al., 2021). We then estimated functional enrichment for each category as a relative risk (i.e., a ratio of proportion of variants) between being in an annotation and fine-mapped (PIP ≤ or PIP > 0.1). That is, a relative risk = (proportion of variants with PIP > 0.1 that are in the annotation) / (proportion of variants with PIP ≤ 0.01 that are in the annotation). 95% confidence intervals are calculated using bootstrapping with 5,000 replicates. We note that, to increase statistical power, we combined pLoF/missense and 5’/3’ UTR into single categories respectively and used a more lenient threshold (PIP > 0.1 vs. > 0.9) compared to our previous analysis (Kanai et al., 2021).

### Gene- and variant-prioritization

To nominate genes using GWAS results, we used the following approach to balance clarity with confidence in the gene assignment.

1. If the locus had a credible set, for each credible set (CS):
  a. Filter to variants in the credible set and retain variants from the CS that are either minimal PIP, coding, or have PIP > 0.7
  b. If the variant has PIP > 0.9 and is a coding variant for a gene, assign that gene to the CS
  c. Otherwise assign genes within 3kb of the variant or, if no genes are within 3kb, assign the nearest gene to the CS
2. If the locus had multiple credible sets and at least one had a variant with PIP > 0.1, we retained assignments only corresponding to variants with PIP > 0.1
3. If the locus did not have a credible set, we assigned the gene with a boundary nearest to the most significant variant in the locus

If a variant is inside a gene body (but is non-coding), we consider that gene to be nearest. For case-only heteroplasmy GWAS, when the same locus was significant across multiple heteroplasmy phenotypes, we performed manual integration to arrive at a set of genes supported by the most compelling genetic evidence across variants for each locus. The SSBP1 locus was particularly complex, so we assign SSBP1 (which harbors the max PIP variant) and provide visualization of the full locus (**Supplementary figure 9K**). We do not use fine-mapping evidence from variants with PIP > 0.1 that are not assigned to a credible set. All assignments were manually reviewed. In all GWAS visualizations, we label the strength of evidence supporting the gene assignment (e.g., if supported by moderate or high-PIP fine-mapped variants, coding variants).

### Colocalization with eQTLs

We conducted colocalization of fine-mapped variants of mtDNA phenotypes and *cis*-eQTL associations from GTEx v8 (Aguet et al., 2020) and eQTL catalogue release 4 (Kerimov et al., 2021) as described previously (Kanai et al., 2021). Briefly, we retrieved fine-mapping results of *cis*-eQTL associations that were fine-mapped using SuSiE (Wang et al., 2020) with covariate-adjusted in-sample dosage LD matrices (Kanai et al., 2021). We then computed a posterior inclusion probability of colocalization for a variant as a product of PIP for GWAS and for *cis*-eQTL (CLPP = PIP_GWAS_ × PIP_*cis*-eQTL_) (Hormozdiari et al., 2016). When displaying colocalization across heteroplasmy traits, we indicate colocalization if we see colocalization PIP > 0.1 for the assigned gene and any variant in the credible set for any tissue and for any heteroplasmy trait.

### Replication of UKB heteroplasmy results in AllofUs

To perform replication analysis in AllofUs, we used LD-independent lead SNPs from all case-only heteroplasmy GWAS originally performed in UKB (**Methods**). We filtered association statistics from AoU (**Methods**) to these lead variants and compared effect sizes when the variants were identified in AoU with MAC > 20. We used Deming regression implemented in the `deming` v1.4 package in R to assess the relationship between effect sizes for these lead SNPs in cross-ancestry meta-analyses in the two biobanks while accounting for standard errors in both (Deming, 1943; Zhou et al., 2022). We coded alleles such that effect sizes were always positive in UKB.

### Assessment of LD with known polymorphic and reference NUMTs

We collated an extensive database of polymorphic and reference NUMT intervals using BLAST, known reference NUMTs (Calabrese et al., 2012; Li et al., 2012), and published polymorphic NUMTs inferred using mate-pair mapping discordance (Dayama et al., 2014; Wei et al., 2022). To search for regions of homology to the mtDNA within the reference nucDNA, we used BLASTn with the GRCh37 reference genome with a word size of 11, an expect threshold of 0.05, short queries enabled, and default values for the other parameters. In total, we obtained 4,736 overlapping reference and polymorphic NUMT intervals. We constructed a 20kb window around each nucDNA NUMT region (10kb up, 10kb down) and then conservatively tested for LD R^2^ > 0.1 between any SNP in the window and each lead variant at genome-wide significance for our UKB case-only heteroplasmy GWAS using in-sample genome-wide EUR LD matrices generated in UKB (*Pan UKBB Initiative*, 2022). All LD values used to examine individual loci in either biobank was computed in-sample – for example, in AoU we computed LD using the post-QC genotype MatrixTable (**Methods**) used for GWAS with the Hail function `hl.ld_matrix()`.

### Multiple alignment of POLG2 protein sequence

POLG2 homologs were detected via best bi-directional BlastP hit (Expect < 1e-3) from human and were aligned via MUSCLE (Edgar, 2004).

