## Supplementary figures and supplementary note for "Nuclear genetic control of mtDNA copy number and heteroplasmy in humans"

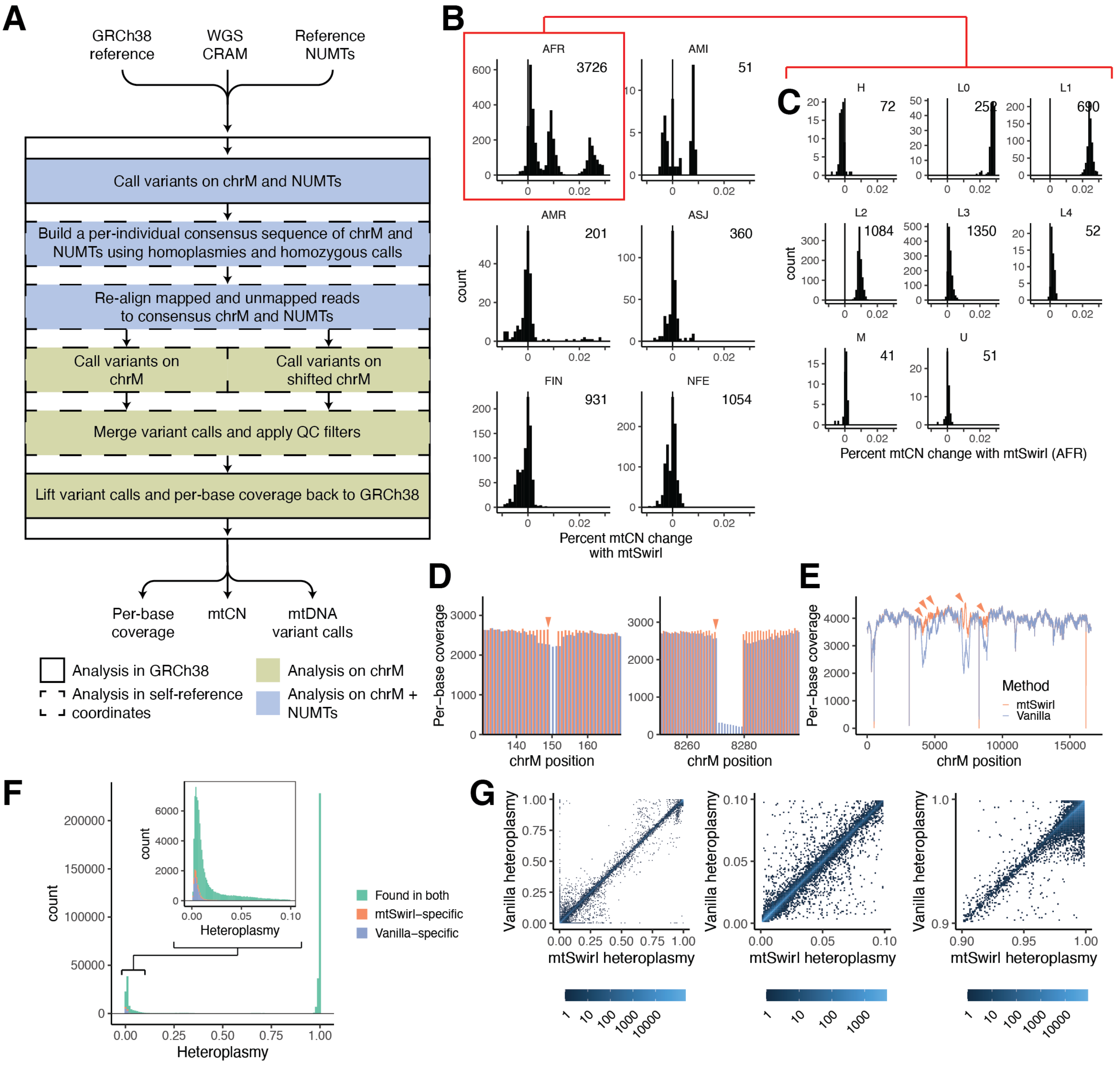


**Supplementary figure 1. Copy number and heteroplasmy estimation improvements using mtSwirl pipeline.**
**A.** Overview of mtSwirl pipeline workflow. Colors represent genomic region analyzed (blue = chrM and NUMTs; yellow = chrM only); border style represents coordinate system (solid = GRCh38; dashed = self-reference coordinates). All output is in GRCh38. **B.** Percent change in mtCN estimated using "vanilla" pipeline versus mtSwirl as a function of inferred nuclear ancestry. **C**. Percent mtCN changes among AFR individuals as a function of mtDNA haplogroup. **D**. Example of per-base coverage improvement with mtSwirl near a homoplasmic indel, likely due to use of mtDNA self-reference sequence. Arrows highlight homoplasmic indels. **E**. Example per-base coverage improvement likely due to reduced mis-mapping to nucDNA. Arrows highlight coverage improvements. **F**. Variant calls found using both pipelines (green), only in mtSwirl (red), and only in "vanilla" (blue). Inset corresponds to zoomed view of low heteroplasmy variants. **G.** 2D histogram showing relationship between heteroplasmy estimates using mtSwirl with "vanilla". Left panel corresponds to overall heteroplasmy space; middle is zoomed to low heteroplasmy variants; right is zoomed to high heteroplasmy variants.

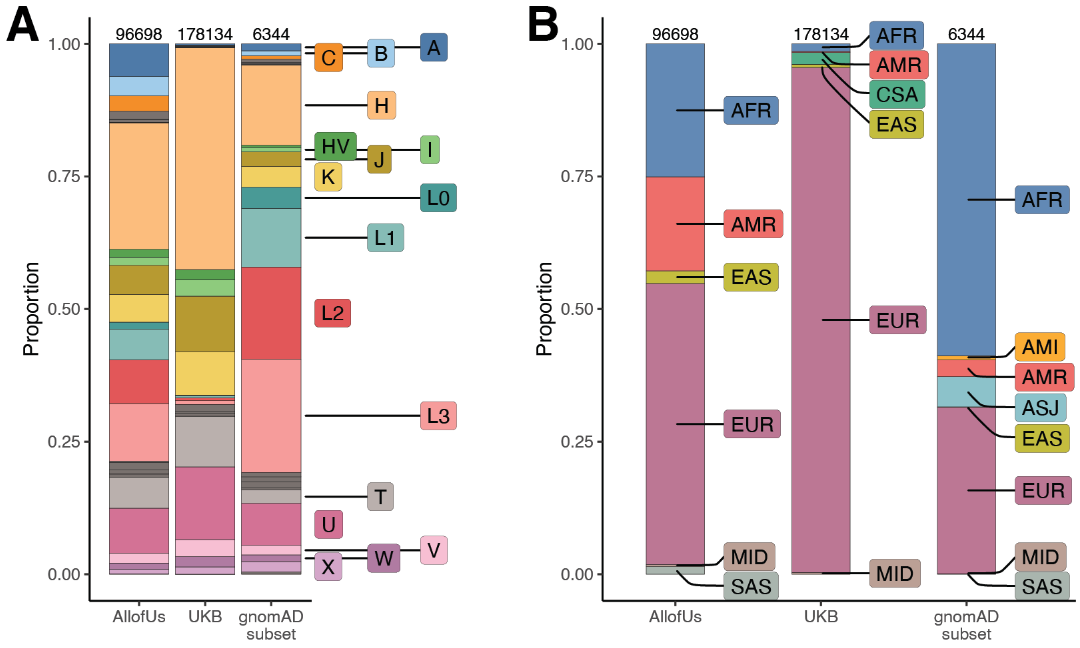


African: L0, L1, L2, L3, L4, L5, L6
Asian: A, B, C, D, E, F, G, M, N, O, P, Q, S, Y, Z
European: H, HV, I, J, K, R, T, U, V, W, X

**Supplementary figure 2. Composition of cohorts used in this study.**

**A.** Top-level haplogroups represented in each analyzed cohort. Labeled haplogroups comprise > 1.5% of the samples in at least one cohort. Haplogroups are mapped to broad ancestral categories in text below the plot with colors corresponding to the colors of the labeled haplogroups. **B.** Inferred nuclear genetic ancestry groups in each analyzed cohort. Ancestry group assignment was completed within each cohort. The NFE and FIN groups in gnomAD were combined under “EUR” for the purposes of this comparison. In both panels, numbers at the top of each bar indicate the number of samples with generated mtDNA callsets passing QC with completed genetic ancestry assignment.

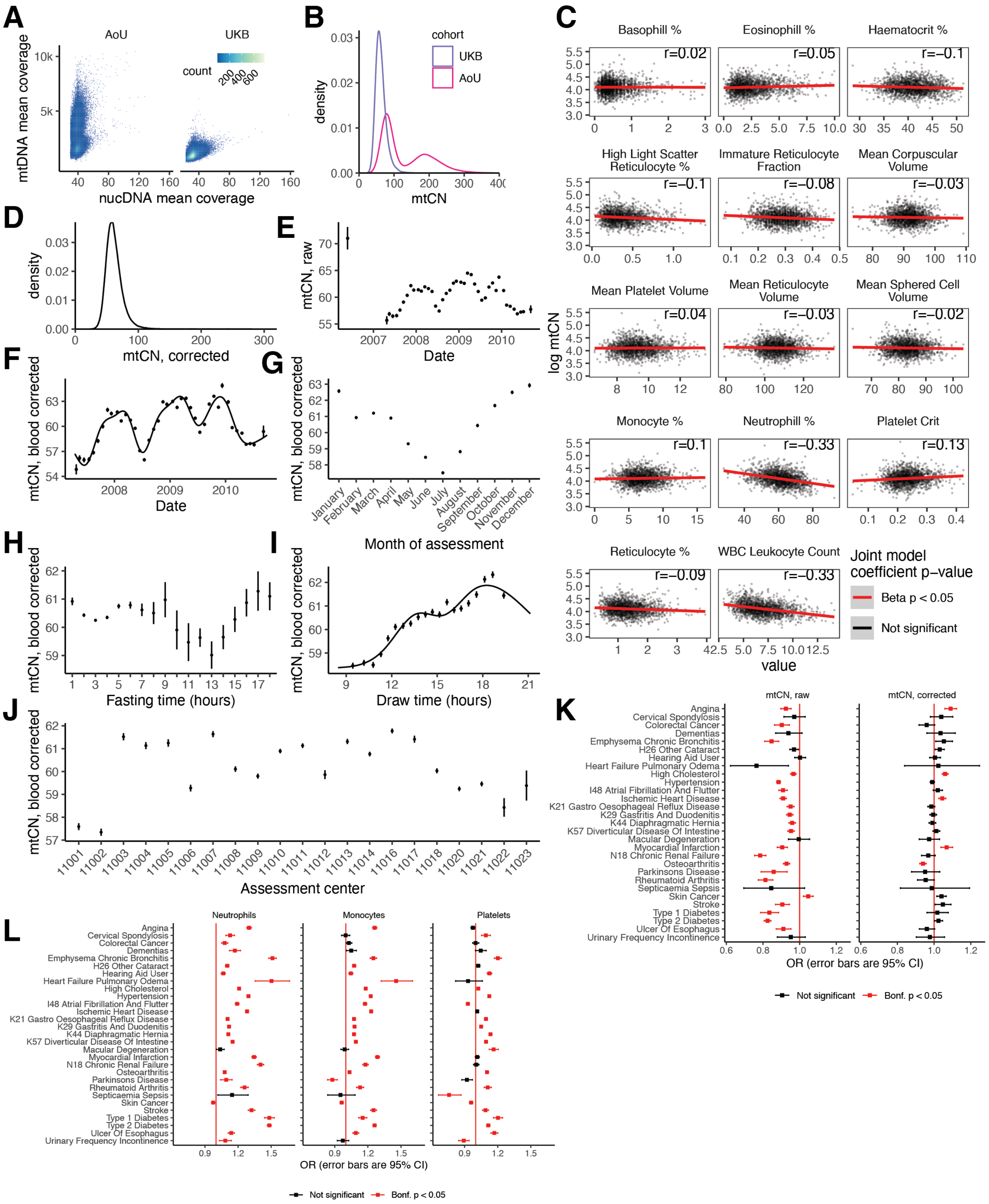


**Supplementary figure 3. mtCN shows substantial correlations with technical and biological covariates.**

**A.** Bivariate mean coverage distributions across nucDNA and mtDNA in AllofUs and UKB. **B.** Distributions of mtCN across AllofUs and UKB. **C**. Correlations between log mtCN and blood cell traits in UKB. Line corresponds to ordinary least squares fit; line color corresponds to coefficient p-value of a joint model regressing log $mtCN_{raw}$ on all blood cell phenotypes. Inset is Pearson correlation coefficient. **D.** Distribution of $mtCN_{corr}$ in UKB. **E.** $mtCN_{raw}$ versus assessment date, binned into months. Pilot month samples are removed from subsequent analyses. **F**. Blood-corrected mtCN as a function of assessment date; line is a natural spline with knots positioned seasonally. **G**. Blood-corrected mtCN as a function of assessment month. **H**. Blood-corrected mtCN as a function of self-reported fasting time. **I**. Blood-corrected mtCN as a function of draw time; line corresponds to natural spline with 5 knots. **J.** Blood-corrected mtCN as a function of assessment center. **K**. OR of raw and corrected mtCN in predicting 29 common diseases in UKB. **L**. OR of top blood cell composition traits in predicting any of 29 common diseases in UKB. For E-L error bars correspond to 1SE. All tests are two-sided.

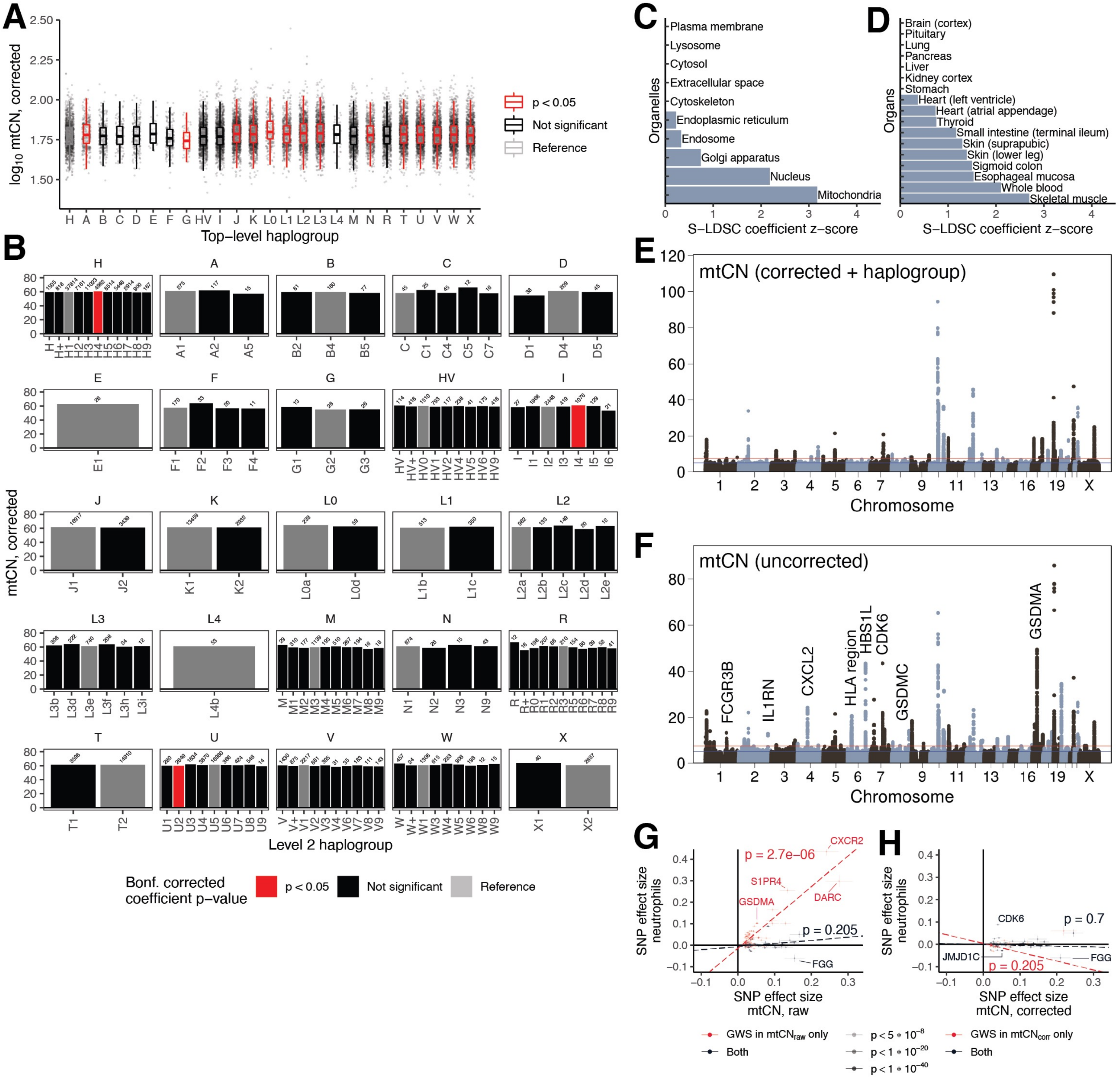


**Supplementary figure 4. The genetic architecture of mtDNA copy number is influenced by blood cell traits but not haplogroup.** **A.** log10 $mtCN_{corr}$ as a function of major top-level haplogroup. Points have been downsampled to at most 1000 per haplogroup. Color represents two-sided regression coefficient p-value from a joint linear model regressing log mtCN onto top-level haplogroup. **B.** Mean $mtCN_{corr}$ as a function of "level 2" haplogroup. Colors correspond to two-sided coefficient p-values for a joint model regressing log mtCN onto level 2 haplogroups within each top-level haplogroup, corrected for multiple testing using the Bonferroni approach across 25 top-level haplogroups. **C**. Enrichment of genome-wide signal near genes annotated to localize to each organelle and **D**. near genes highly expressed in each tissue. **E**. GWAS Manhattan plot of $mtCN_{corr}$ additionally corrected for top-level haplogroup. **F**. GWAS Manhattan plot of $mtCN_{raw}$. Labels indicate genes proximal to a non-exhaustive set of selected loci with substantially less-significant p-values in the corrected analysis. **G**. Correlation between effect sizes for lead SNPs detected for raw mtCN between $mtCN_{raw}$ and neutrophil count. **H**. Correlation between effect sizes for lead SNPs detected for $mtCN_{corr}$ between $mtCN_{corr}$ and neutrophil count. In panels G and H, error bars represent +/- beta SE, dotted line corresponds to weighted least squared regression line; inset corresponds to regression p-value. Regression fits were performed separately for loci genome-wide significant for both $mtCN_{raw}$ and $mtCN_{corr}$ (black) and for loci specific to each (red).

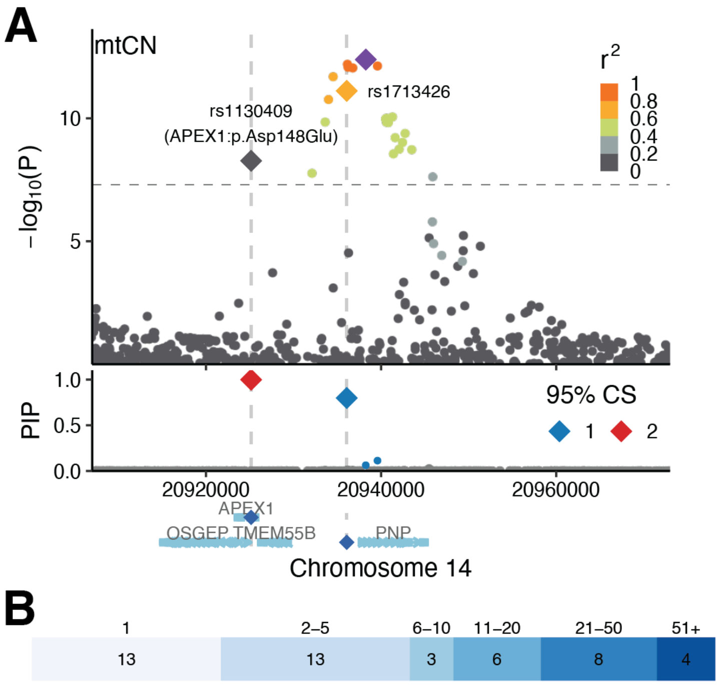


**Supplementary figure 5. Chromosome 14 locus shows likely causal variants in two adjacent genes.**
**A.** Upper panel shows UKB GWAS meta-analysis p-values at the chromosome 14 locus. Middle panel shows variants in the two 95% credible sets identified at this locus, with large diamonds corresponding to the highest PIP variants in each credible set. Bottom panel shows protein-coding gene annotations at this locus. Variant overlapping APEX1 is a missense variant in APEX1. **B.** Distribution of sizes of credible sets identified via fine-mapping for $mtCN_{corr}$. Numbers atop shaded region correspond to size of CS; numbers within shaded region corresponds to the count of credible sets of that size.

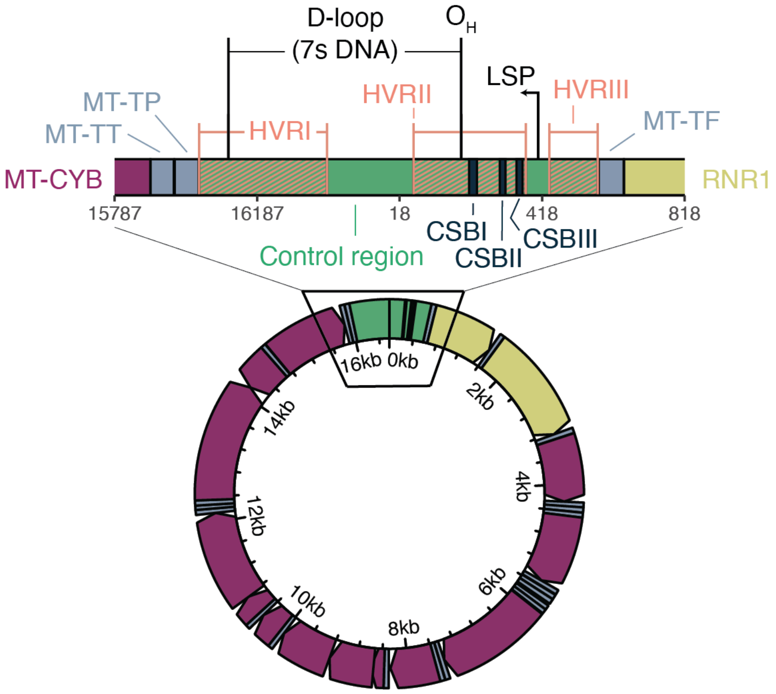


**Supplementary figure 6. Organization of the mtDNA control region.**

Colors indicate annotation type. Yellow indicates rRNA gene; steel represents tRNA gene; purple represents coding genes; green represents the control region; and midnight represents the conserved sequence boxes (CSB). Salmon stripe pattern represents the hyper-variable regions (HVR). The mtDNA D-loop refers to the region within the control region often showing triple-stranded DNA due to the persistence of the 7s DNA. Annotations are oriented with the rCRS reference genome.

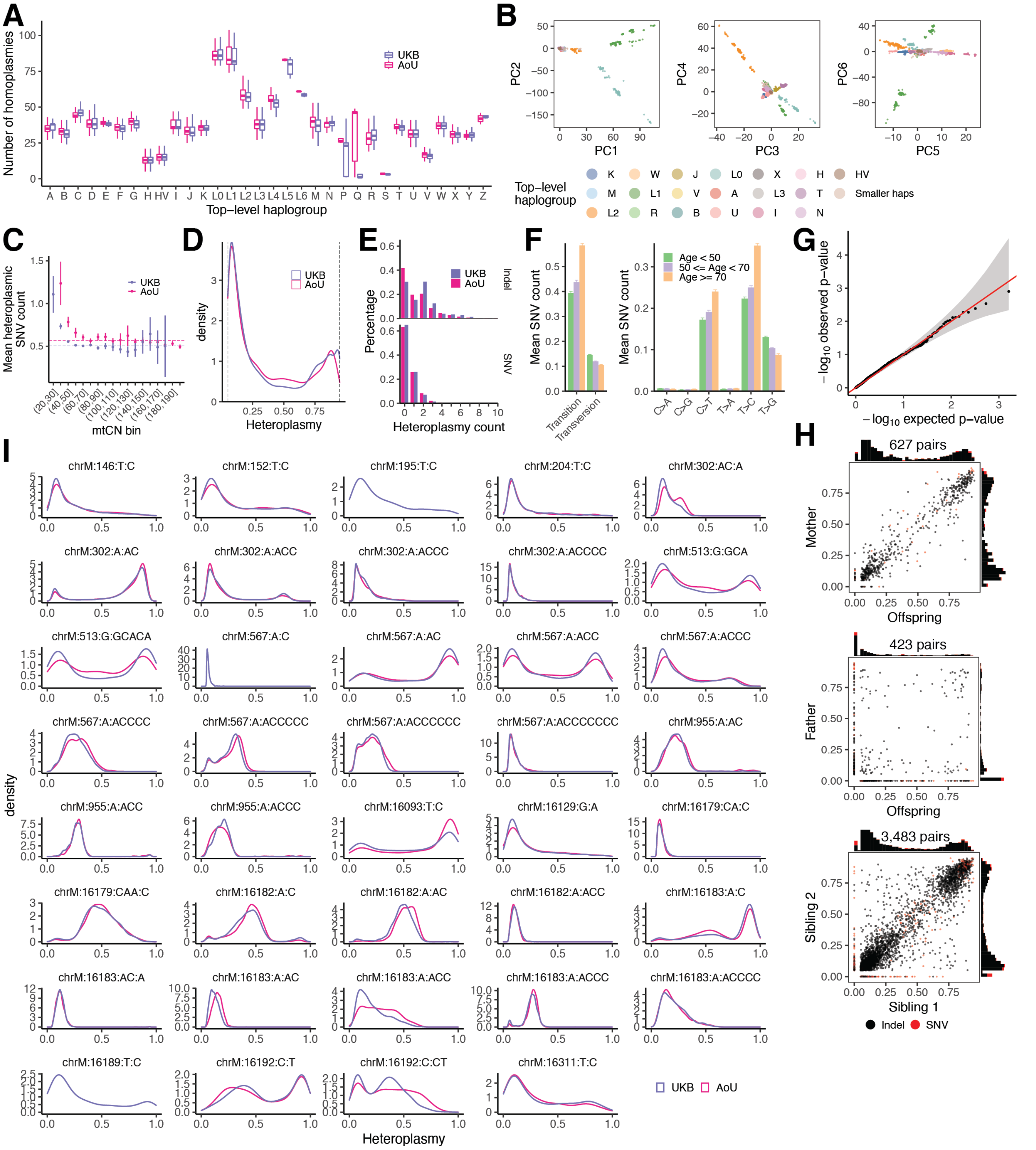


**Supplementary figure 7. Overview of mtDNA variation across ~274,000 individuals.**

**A.** Box-and-whisker plots of homoplasmies per mtDNA haplogroup. Colors correspond to biobank. Outliers are suppressed to prevent visualizing AoU individual-level data. **B.** Projection of UKB samples into mtDNA PC space computed using homoplasmies (MAF > 0.001). **C**. Heteroplasmic SNV count as a function of mtCN in UKB and AllofUs. Dotted lines correspond to mean number of heteroplasmic SNVs per person for individuals with mtCN > 50. Plot is truncated at mtCN < 200 for viewability. **D**. Heteroplasmy distributions restricted to between 0.05 and 0.95 across UKB and AoU. **E**. Histogram of heteroplasmy counts per person for indels (top) and SNVs (bottom). **F**. Mean SNV count identified per-person in AoU as a function of variant type and age group. Error bars are +/- 1SE. **G**. Quantile-quantile plot of p-values from logistic regression tests predicting case/control status of 29 common diseases in UKB using each of 39 common case-only heteroplasmies (see panel I). Ribbon corresponds to 95% CI around expectation. **H**. Heteroplasmy correlations for 39 common heteroplasmies (see panel I) in mother-offspring (top), father-offspring (middle), and sibling-sibling pairs (bottom). **I**. Case-only heteroplasmy distributions of 39 variants detected in >500 UKB samples.

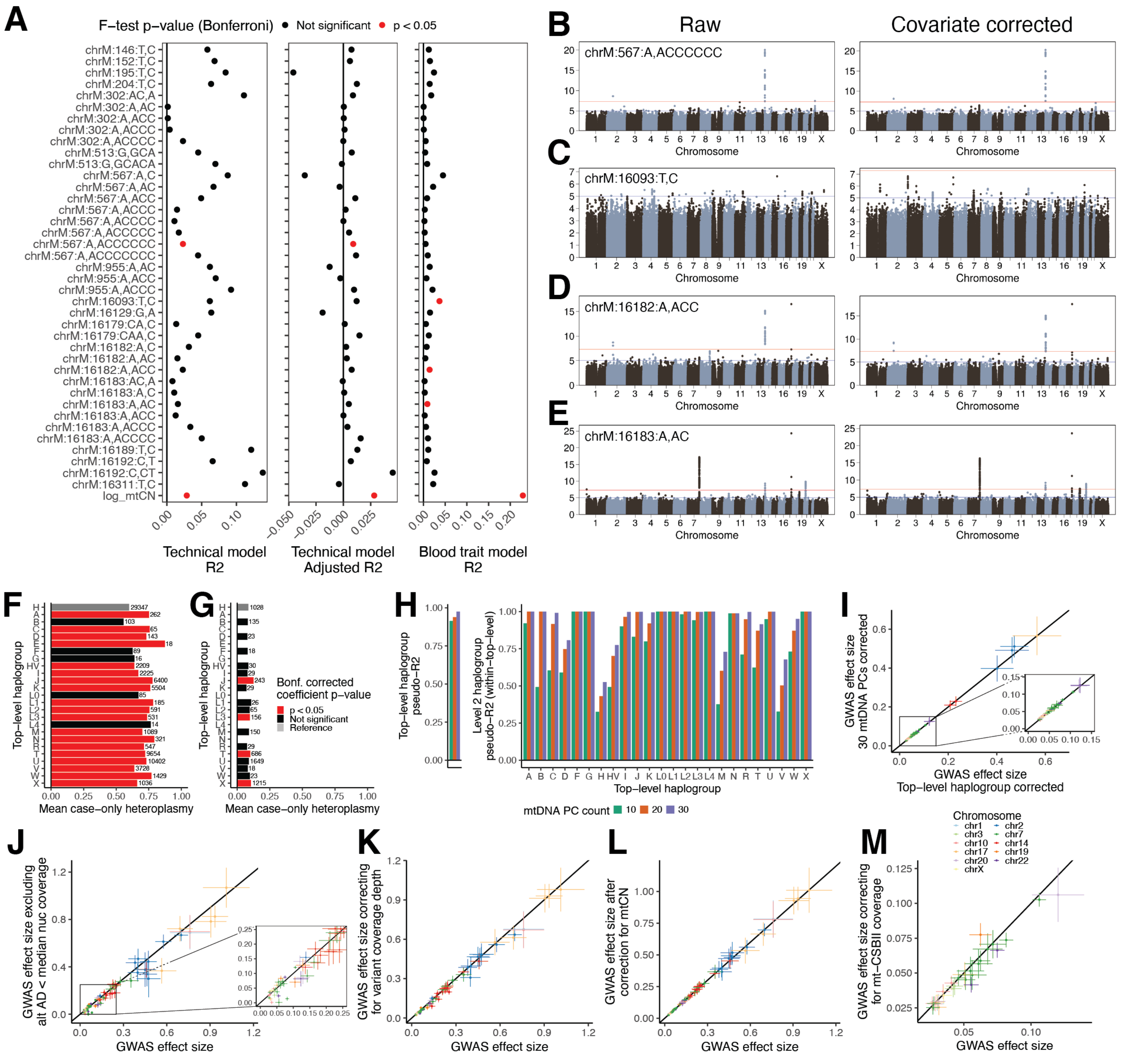


**Supplementary figure 8. mtDNA heteroplasmy estimates and genetic associations are robust to potential confounders.**

**A.** $R^2$ and adjusted $R^2$ for technical covariate model and $R^2$ for blood trait model for common mtDNA heteroplasmies and log $mtCN_{raw}$. Color corresponds to model F-test p-value < 0.05 (df=N-14 for blood, N-67 for technical; N in **Supplementary table 1**) after Bonferroni correction. Sensitivity analyses of the GWAS for **B.** chrM:567:A,ACCCCCCC before and after technical covariate correction, **C.** chrM:16093:T,C before and after blood trait correction **D.** chrM:16182:A,ACC before and after blood trait correction **E.** chrM:16183:A,AC before and after blood trait correction. Mean case-only heteroplasmy as a function of top-level haplogroup for **F.** chrM:302:A,AC and **G.** chrM:16179:CA,C. Bar color corresponds to two-sided coefficient p-value for the regression of heteroplasmy onto top-level haplogroup, Bonferroni corrected for 39 tested heteroplasmies. **H**. McFadden's pseudo-$R^2$ for a multinomial model of top-level haplogroup versus mtDNA PCs (left) and "level 2" haplogroup versus mtDNA PCs within each top-level haplogroup. **I**. GWAS lead SNP effect size estimate correlation when correcting for 30 mtDNA PCs vs correcting for only top-level haplogroup for selected variants showing high haplogroup heterogeneity (302:A,AC; 302:A,ACC; 302:A,ACCCC; 567:A,ACCCCCC; 955:A,ACC; 16179:CA,C; 16183:A,C). GWAS lead SNP effect size estimate correlation between case-only GWAS at baseline and **J**. GWAS after removing heteroplasmy calls supported by allele depth < median nuclear coverage, **K.** GWAS after correcting for variant coverage depth, **L.** GWAS after correcting for mtCN, **M**. length heteroplasmy GWAS after correcting for CSBII median coverage. For panels I-M, colors correspond to nuclear chromosome, points correspond to lead SNPs from baseline case-only GWAS including top-level haplogroup covariates, and error bars represent +/- 1 SE.

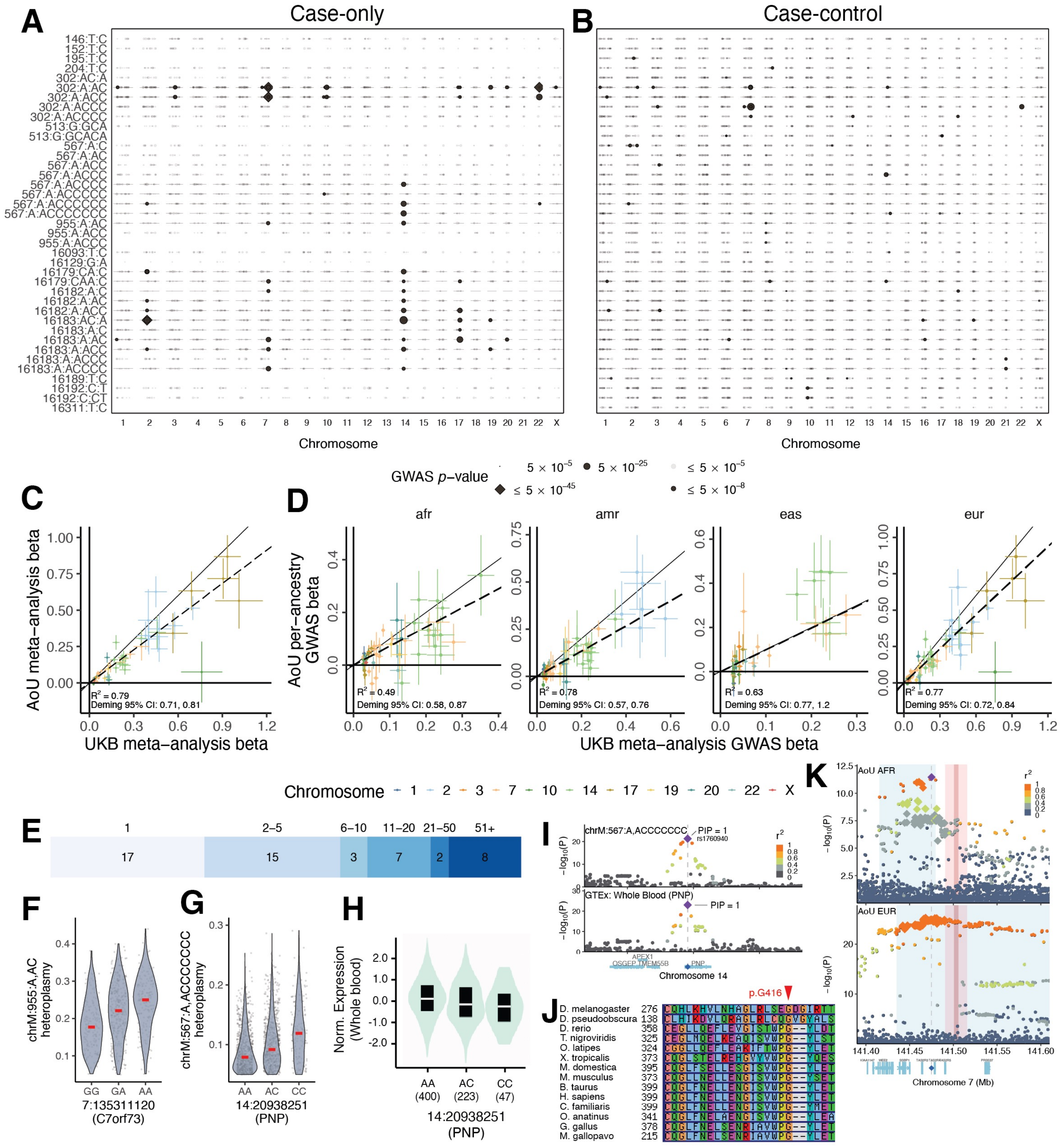


**Supplementary figure 9. The full landscape of nuclear genetic associations to common mtDNA heteroplasmies.**

**A.** Lead SNP p-values across all 39 tested case-only mtDNA heteroplasmies in the style of Figure 5c. **B.** Lead SNP p-values across all 39 tested mtDNA heteroplasmies when coded as case-control phenotypes. **C**. Replication of lead SNP-variant pairs tested in both the UKB meta-analysis and AllofUs meta-analysis for case-only heteroplasmy. Error bars correspond to 1SE; colors correspond to nuclear chromosome. **D**. Replication of lead SNP-variant pairs tested in the UKB meta-analysis with each AoU continental ancestry group. Error bars correspond to +/- 1 effect size SE; colors correspond to nuclear chromosome. **E.** The distribution of 95% credible set sizes from all heteroplasmy GWAS. Numbers atop shaded region correspond to size of CS; numbers within shaded region corresponds to the count of credible sets of that size. **F**. chrM:955:A,AC heteroplasmy as a function of lead SNP genotype near C7orf73. **G.** chrM:567:A,ACCCCCCC heteroplasmy as a function of highest PIP SNP genotype in PNP. **H**. Whole blood PNP expression as a function of the same highest PIP SNP genotype in GTEx. **I**. Colocalization between chrM:16183:AC,A at the PNP locus and PNP eQTL in whole blood. **J.** Multiple sequence alignment across vertebrates of best bidirectional hits for POLG2 (BLASTP E<1e-3) displayed with ClustalW colors with effect of putative causal variant labeled. **K**. GWAS results in AoU for AFR and EUR in the vicinity of SSBP1 for chrM:302:A,AC. Large points correspond to 95% CS from UKB meta-analysis, blue ribbon is region with LD R2 > 0.8 to lead SNP, dark red ribbon is a reference NUMT, light red ribbon is a 20kb window around the reference NUMT.

**Supplementary note 1 – full description of mtSwirl pipeline**

Upon ingestion of the aligned whole genome read file, we use GATK v4.2.6.0 PrintReads to quickly subset from the CRAM / BAM file to a much smaller file containing reads mapping to either the mtDNA, any intervals provided as reference NUMTs, or any other unmapped reads. Any paired-end reads which span the boundary of a reference NUMT interval are removed; we extend each NUMT interval by 500 bases prior to input to avoid any significant loss of coverage in the body of the interval.

We use these reads to perform the first round of variant calling in the NUMT regions and on the mtDNA, implementing filters to ensure only the highest quality variant calls are considered for self-reference construction. For nuclear DNA variant calls we use GATK best practices for single-sample variant calling with HaplotypeCaller. For SNVs we remove calls with QD < 2.0, QUAL < 30, SOR > 3.0, FS > 60.0, MQ < 40.0, MQRankSum < -12.5, ReadPosRankSum < -9.0, and a read depth of < 10. For INDELs we remove calls with QD < 2.0, QUAL < 30.0, FS > 200.0, SOR > 10.0, ReadPosRankSum < -20.0, and a read depth of < 10. For the mtDNA we run Mutect2 in mitochondria mode using settings as described previously (Larrichia et al. 2022 Genome Res), removing any variants flagged by FilterMutectCalls or overlapping regions previously described as artifact-prone (300-302; 309-310; 315-316; 3106-3107; 16181-16182). We use Haplochecker (Weissensteiner et al., 2021) Click or tap here to enter text.to estimate mitochondrial contamination and remove mtDNA variant calls potentially influenced by contaminated reads using FilterMutectCalls.

First round variant calls are next used to construct a self-reference sequence for each sample. Specifically, we filter all high-quality variant calls to homoplasmies (HL > 0.95, mtDNA) or homozygous alternate (NUMT regions) calls. In rare instances, we observed homoplasmic variant calls on the mtDNA that were overlapping (e.g., a homoplasmic chrM:513:GCACA,G and a homoplasmic chrM:513:G,C) – we remove overlapping variants and flag these samples. In NUMT regions, rare instances of overlapping variants or variants overlapping interval boundaries are skipped. The consensus sequence is constructed with bcftools v1.16 consensus (Danecek et al., 2021) using the final high quality homoplasmic and homozygous variants list for the mtDNA and NUMT regions respectively. The pipeline also produces a shifted version of the mtDNA consensus sequence as done previously (Laricchia et al., 2022) to improve read mapping to the ends of the circular chromosome.

To facilitate read mapping, we reproduce several reference files in self-reference coordinates including the coordinates of the mtDNA control region, the length of the mtDNA, any blacklisted intervals, and chain files to allow for coordinate mapping between the self-reference genome and GRCh38. Liftover of coordinates was performed using UCSC liftOver tools (https://github.com/ucscGenomeBrowser/kent). We also produce a VCF containing the set of mtDNA high quality homoplasmies used for consensus construction in self-reference coordinates – these are used to produce “force-calls” using Mutect2.

For the second round of variant calling, we produce an unmapped SAM file and then use bwa mem (with parameters -K 100000000 -p -v3) to map reads onto the self-reference and shifted self-reference sequences containing the mtDNA and NUMT regions. Picard MarkDuplicates and SortSam are subsequently used to pre-process the set of mapped reads prior to extracting only reads mapping to chrM. Mutect2 is then used in mitochondria mode to call variants on the mtDNA and shifted mtDNA using the same parameters as in the first round of variant calling, including the "force-call" variants to ensure that variant call data is obtained from sites that are homoplasmic relative to GRCh38 and incorporated into the consensus sequence. Calls from the control region as obtained from the shifted mtDNA are merged with calls elsewhere on the mtDNA and artifactual variant calls, including those likely influenced by contamination (as estimated during the first round of variant calling), are filtered using FilterMutectCalls.

The final step of the pipeline involves returning variant calls and per-base coverage estimates to GRCh38 for use in population-scale analyses. We do this via a two-step approach: first we use Picard LiftoverVcf to move "simple" variants back to GRCh38. Many variants fail this step – all force-called variants, for example, are former heteroplasmies in GRCh38 that were incorporated into the self-reference and thus have a reference allele that does not match the GRCh38 sequence. We developed a custom Liftover pipeline which systematically handles all major cases of Liftover failure, including reference allele mismatch, variants present within homoplasmic insertions relative to GRCh38, deletions spanning sites which differ between the consensus sequence and GRCh38, insertions which share a position with a homoplasmic deletion relative to GRCh38. Across >4,000,000 variant calls across 199,919 samples in UKB, we observed only 73 variants across 64 samples that were "failed" by both Picard LiftoverVcf and our custom Liftover pipeline. For per-base coverage, we use kent LiftOverTools to lift self-reference coverage estimates back to GRCh38. In instances where deletions relative to GRCh38 were incorporated into the self-reference, the allele depth of the reference allele of the homoplasmic variant call (relative to GRCh38) of these deletions is used as the per-base coverage in these gaps.

### Supplementary note 2 – analysis of technical and biological covariates in mtDNA phenotypes

We closely analyzed mtDNA phenotypes for associations with technical and biological covariates, with the former including assessment center, time of day of blood draw, fasting time, month of year, and assessment date, and the latter including blood cell composition and mtDNA population structure (i.e., haplogroup).

We started by assessing the relationship between mtDNA phenotypes and blood cell type percentages and mean blood cell volumes (**Methods**). All analyses were performed with log(mtCN) as the response variable. We observed strong linear relationships between log(mtCN) and most tested blood phenotypes (**Supplementary figure 3C**), with a joint linear model of tested blood phenotypes explaining close to 23% of the observed variance in log(mtCN) (**Figure 1A**). In contrast, on testing heteroplasmies with a joint linear model of blood cell phenotypes, we observed only 3 heteroplasmies with evidence of having at least one nonzero linear model coefficient (F-test $p < 0.05$ after Bonferroni correction; **Supplementary figure 8A**). We ran GWAS of these three case-only heteroplasmies (chrM:16093:T,C, chrM:16182:A,ACC, chrM:16183:A,C) with and without blood cell phenotype correction and observed very little visible difference in spectrum of genome-wide significant hits (**Supplementary figure 8C-8E**). Based on these results, we implemented corrections for blood cell phenotypes in analyses of mtCN only.

We next investigated time of day of blood draw, fasting time, assessment date, and assessment center as technical covariates (see **Methods** for model specification). We found that UKB samples processed in 2006 (N = 51) showed a significantly different average mtCN compared to other periods (**Supplementary figure 3E**). Given the small sample size of this period, the fact that this was a pilot, and the different average mtCN, we exclude samples collected in 2006 from all subsequent analyses. For subsequent analyses of technical variables on mtCN, we pre-correct mtCN by obtaining residuals from the model: $\log mtCN \sim blood\ cell\ variables$. Blood-corrected mtCN showed seasonal variability (**Supplementary figure 3F, 3G**), differences across fasting time (**Supplementary figure 3H**), differences as a function of the time of day at which blood was drawn (**Supplementary figure 3I**), and evidence of heterogeneity across assessment center (**Supplementary figure 3J**). A model consisting of blood draw time, fasting time, assessment date, and assessment center explained ~3% of the variation in $mtCN_{raw}$ (**Figure 1A**) with significant evidence in support of non-zero coefficients (F-test p-value < 2.2e-16). Similar patterns were not observed for case-only heteroplasmy measures across all tested variants – the same technical covariate model showed variable $R^2$ estimates across the case-only mtDNA heteroplasmies (**Supplementary figure 8A**), and failed to show evidence in support of any coefficients significantly different from zero in all cases (F-test p-value > 0.05 after Bonferroni correction) other than the variant chrM:567:A,ACCCCCC. Adjusted $R^2$ estimates showed greatly reduced magnitude for heteroplasmy traits while remaining stable around ~0.03 for mtCN (**Supplementary figure 8A**). We conducted case-only GWAS (**Methods**) for chrM:567:A,ACCCCCC with and without correction for technical covariates and found minimal-to-no changes in the prevailing genome-wide architecture with correction (**Supplementary figure 8B**). Based on these results, we implemented corrections for these technical covariates in analyses of mtCN only.

We assessed the impact of mtDNA population structure via haplogroup and homoplasmic mtDNA variation on $mtCN_{corr}$. We observed that top-level haplogroup showed very small but significant associations with $mtCN_{corr}$ (**Supplementary figure 4A**), with top-level haplogroup explaining < 0.5% of the variance in $mtCN_{corr}$. As haplogroups form a nested tree structure of ancestral variation, we next asked how much $mtCN_{corr}$ varied between "level 2" haplogroups within each top-level haplogroup. We found very little evidence of mean differences in log(mtCN) across level 2 haplogroup within each top-level haplogroup (**Supplementary figure 4B**). As a sensitivity analysis, we repeated the $mtCN_{corr}$ GWAS including top-level haplogroup indicator variables as covariates in the GWAS model and observed no observable change to the genome-wide architecture (**Figure 1D**, **Supplementary figure 4E**). Thus, we do not correct mtCN for haplogroup in our analyses.

Finally, we examined mtDNA population structure correlations with tested mtDNA heteroplasmies. In contrast with $mtCN_{corr}$, we observed many significant associations of top-level haplogroup on case-only heteroplasmy levels (**Supplementary figure 8F, 8G**) and thus include top-level haplogroup indicator variables in all heteroplasmy GWAS (case-only and case/control,

**Methods**). Given the tree structure of haplogroups, we wondered if "deeper" haplogroups also contributed substantial variance to case-only heteroplasmy levels. To avoid inclusion of a prohibitively large set of haplogroup indicator variables, we instead computed the first 30 principal components (PCs) using all QC-pass homoplasmies with MAF > 0.001 in UKB (**Methods**). We found strong separation of samples belonging to different top-level haplogroup (**Supplementary figure 7B, 8H**) as a function of the top PCs, and observed that top 30 PCs have substantial predictive power in assigning samples within each top-level haplogroup to the appropriate "level 2" haplogroup (**Supplementary figure 8H**), indicating that the top 30 mtDNA PCs can effectively replace inclusion of dummy variables for top-level haplogroup while also accounting for deeper levels of haplogroup structure. To test the influence of deeper haplogroup structure on the identified genome-wide architecture of case-only heteroplasmy, we repeated GWAS for 7 representative mtDNA heteroplasmies (chrM:302:A,AC; chrM:302:A,ACC; chrM:302:A,ACCC; chrM:567:A,ACCCCCC; chrM:955:A,ACC; chrM:16179:CA,C; chrM:16183:A,C) now including 30 mtDNA PCs as covariates instead of top-level haplogroup in the GWAS model. We found no evidence of changes in estimated effect sizes for lead SNPs when using 30 mtDNA PCs instead of top-level haplogroup (**Supplementary figure 8I**). Thus, as including mtDNA PCs did not appear to meaningfully influence genetic associations, we include only top-level haplogroup dummy variables in the GWAS model for all heteroplasmy traits.

The final mtCN correction model from which residuals were obtained was:

$$\log mtCN \sim ns(blood\ draw\ time, 5) + assessment\ center + fasting\ time \\ + ns(assessment\ date, SEASONAL\ KNOTS) + month\ of\ assessment \\ + blood\ cell\ variables$$

where the blood cell variables used were hematocrit percentage, platelet crit, monocyte percentage, basophil percentage, eosinophil percentage, neutrophil percentage, reticulocyte percentage, high light scatter reticulocyte percentage, immature reticulocyte fraction, mean corpuscular volume, mean reticulocyte volume, mean sphered cell volume, and mean platelet thrombocyte volume.

For our heteroplasmy phenotypes, we do not apply any corrections and include the usual covariates as well as top-level haplogroup as covariates in the GWAS model (**Methods**). Although we believe our results indicate that heteroplasmy is more robust to technical variation than mtCN, we do have significantly fewer measurements of the common heteroplasmies than mtCN, reducing our power to resolve very subtle influences of these covariates on variant heteroplasmy.

**Supplementary note 3 – use of AllofUs data for mtCN and heteroplasmy analyses**

We quantified all mtDNA phenotypes in both UKB and AoU (**Methods**). Through extensive testing in UKB, we found that technical and biological covariates (e.g., blood cell composition at time of blood draw, date of blood draw, assessment center) had profound impacts on mtCN measurements (**Supplementary note 2, Supplementary figures 3, 4**) with covariates collectively explaining over 26% of the variance in mtCN (**Figure 1A**). Blood cell composition at the time of blood sampling has a particularly large impact on both phenotypic correlations (**Figure 1F, Supplementary figure 3K, 3L**) and genetic locus discovery (**Supplementary figure 4F**) in UKB. Unfortunately, these variables were unavailable in AoU. We observed a suspicious bimodal distribution of mtDNA coverage and mtCN in AoU (**Supplementary figure 3A, 3B**); corrections for assessment state and sequencing center did not ameliorate this pattern. The two modes show vastly different mtCN estimates (~80 per cell versus ~200 per cell), which we believe is too large to be driven by typical biological differences in the population and is more likely to be driven by technical factors (e.g., differences in the fraction of blood used, such as buffy coat versus whole blood, or differences in DNA extraction method). Information on these factors in AoU was unavailable. Thus, we restricted our analysis of mtCN to UKB only to avoid adding bias from uncorrected analyses in AoU.

Importantly, our analysis in UKB showed that heteroplasmy measurements were quite robust to these technical variables (**Supplementary note 2**), with haplogroup being the major covariate associated with heteroplasmy levels (**Supplementary note 2**). We do not include technical or blood composition covariates in our heteroplasmy GWAS or in phenotypic correlation analyses as we observed that very few heteroplasmy traits showed significant associations with any tested covariate (**Supplementary figure 8A**). We performed sensitivity analyses including these covariates in the GWAS model for the four heteroplasmy traits that showed a significant phenotypic association with technical and/or blood composition traits and found no noticeable change in the identified genetic architecture (**Supplementary figure 8B-8E**). Thus, we proceeded with use of AoU mtDNA heteroplasmy calls as replication for UKB, including inferred top-level haplogroup as a covariate along with age, sex, $age^2$, interactions, and PCs in AoU genetic analyses (**Methods**). Importantly, this is the same GWAS model used in UKB. Encouragingly, we observed excellent replication of UKB cross-ancestry meta-analysis lead SNP effect sizes in the AoU cross-ancestry meta-analysis (**Supplementary figure 9C**) and even in GWAS including individual ancestry groups in AoU (**Supplementary figure 9D**), with some effect size attenuation seen consistent with Winner's curse.

**Supplementary note 4 – improvement of residual stratification by computing per-ancestry PCs in AllofUs**

As a sanity check, we extracted four well-powered, well-measured phenotypes in AoU for genetic analysis. We used a SQL query to obtain values corresponding to the measurement concept IDs: 903133, 3004501, 903115, 903118, corresponding to body height, blood glucose, systolic blood pressure, and diastolic blood pressure. We extracted the measurement date for each of these values and filtered to the latest measurement per individual for each phenotype. Because each phenotype was potentially measured at a different date and time, we constructed an age covariate specific to each measurement, corresponding to the age of each individual at the date of measurement of each phenotype. To accomplish this we estimated age as the difference in years between the year of measurement and the year of birth, producing an "age" covariate that was specific for each sample and phenotype. We used the GWAS model as described in **Methods** with the hl.linear_regression_rows() method: age, sex, age*sex, $age^2$, $age^2$*sex, and the 16 PCs initially provided using PCA over all samples using high quality markers on chromosome 20 and

21, where all covariates incorporating age used the computed value corresponding to the estimated age of the participant at the time of measurement of each phenotype.

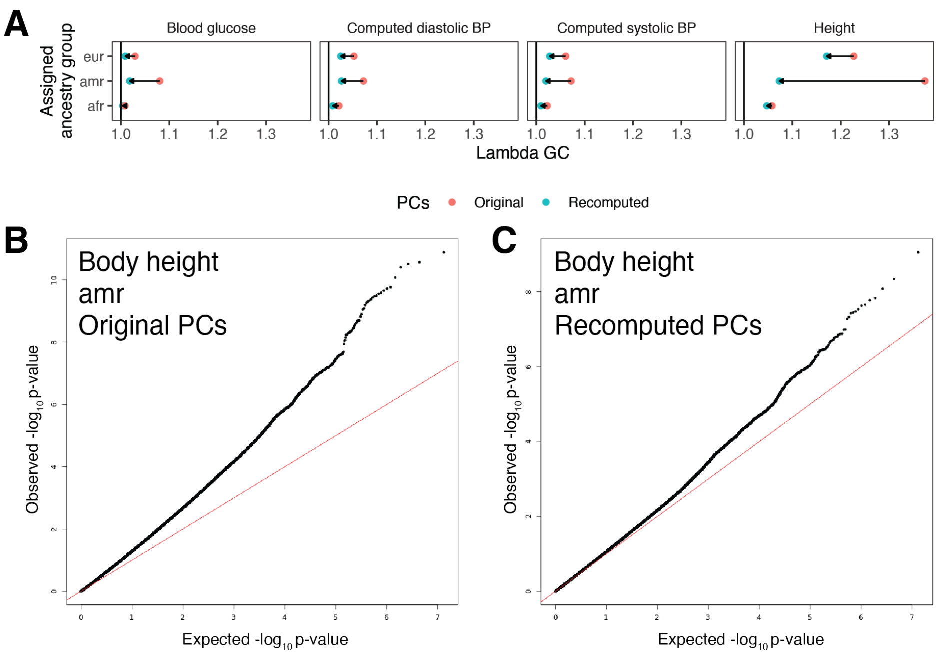


**Supplementary note figure S1**. Improvement of population stratification with recomputed PCs. **A**. Lambda GC values obtained from GWAS summary statistics for 4 positive control phenotypes in AllofUs using original and recomputed PCs as covariates in linear regression GWAS for the three largest ancestry groups. **B**. Quantile-quantile plot of GWAS p-values for body height in the AMR group using original PCs. Red line is the y=x line. **C.** Quantile-quantile plot of GWAS p-values for body height in the AMR group using recomputed PCs.

To our surprise, we noticed evidence of population stratification for several phenotypes as elevated lambda GC (**Supplementary note figure S1A**). GWAS in individuals assigned the AMR group seemed particularly susceptible, with a higher lambda GC than EUR seen for all 4 tested traits despite less than half the sample size. The most extreme example of this was for height, which showed a lambda GC of ~1.37 clearly suggestive for uncorrected stratification (**Supplementary note figure S1B**).

To address this, we recomputed PCs within each assigned population group using markers distributed throughout the whole genome. We imported the AllofUs WGS variant calls in Hail, splitting multi-allelic sites and removing samples and variants failing any QC filters provided by the DRC (**Methods**). We next obtained the high-quality set of variants used for PCA in gnomAD v3 prior to LD-pruning. In brief, autosomal SNVs with no evidence of deviation from Hardy-Weinberg equilibrium within gnomAD (P > 1e-8), an exome call-rate of 0.99, and a gnomAD v3 minimum AC of 10 located outside of segmental duplications and low complexity regions and inside high-coverage exome intervals were obtained. See https://github.com/Nealelab/ccdg_qc/blob/master/scripts/pca_variant_filter.py for more

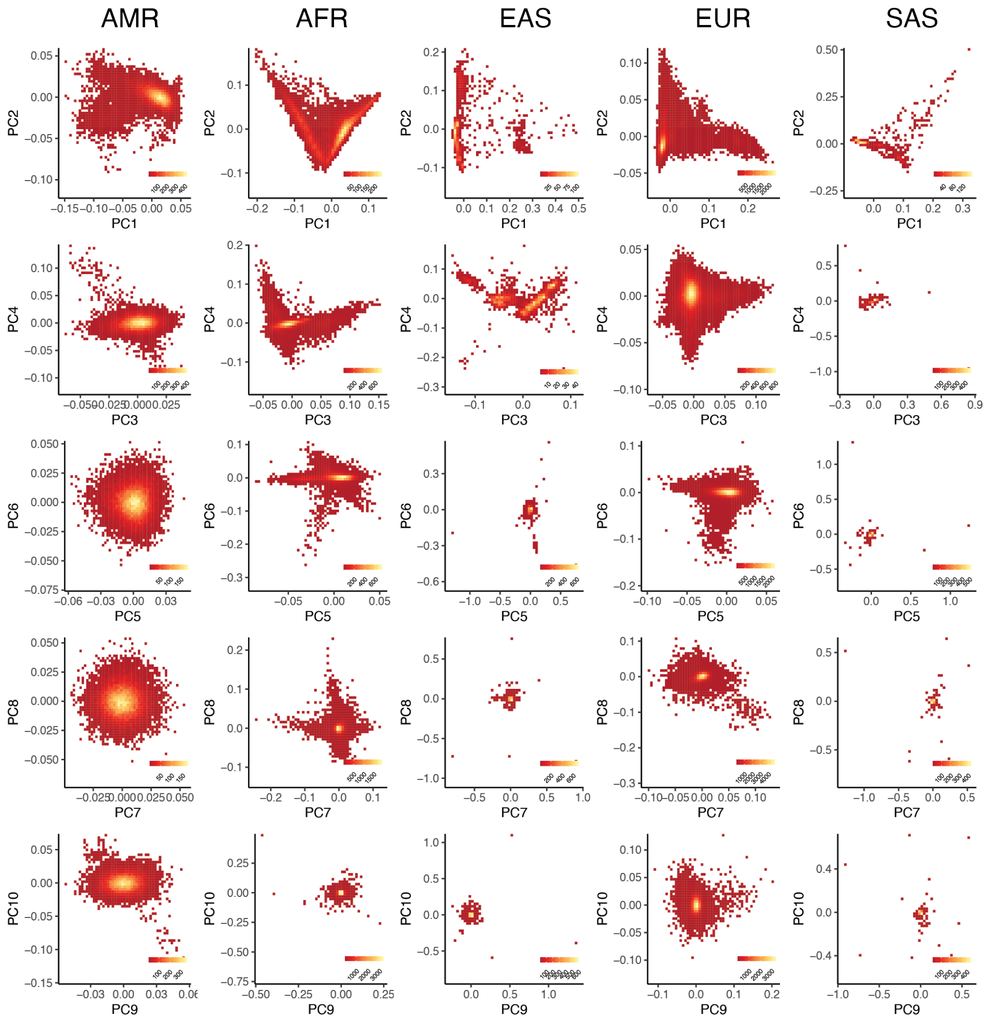


**Supplementary note figure S2**. PCA biplots up to PC10 using PCs recomputed within each major assigned ancestry group in AllofUs. Plots are 2D histograms with per-plot legend corresponding to density scale for each plot.

details on the construction of the high-quality variant set. Of the 259,482 such variants identified, 258,457 variants were found in the AllofUs call-set after QC. Within each assigned ancestry, we filtered to high-quality variants with minor allele frequency > 0.001 and then ran linkage disequilibrium (LD) pruning using hl.ld_prune() with an $r^2$ threshold of 0.1 to identify independent high-quality markers for each ancestry group. Samples corresponding to MID were excluded given the small sample size of these samples. After filtering and pruning, 137,394 variants were included for PCA in at least 1 ancestry group. PCA was performed similarly as for UKB (**Methods**) – for each assigned ancestry group, PCs were produced using LD-pruned high-quality variants for that ancestry among unrelated individuals using the hl.hwe_normalized_pca() method, and related samples were subsequently projected into this PC space. The first 20 PCs were retained for all samples for use as covariates for GWAS. We visualized the first 10 PCs in biplots for each of the five analyzed ancestry groups (**Supplementary note figure S2**), observing generally expected population structure captured by the PCs with few examples of extreme ancestry outliers.

We reran GWAS for our four selected positive control phenotypes using all 20 newly recomputed per-population PCs as covariates and observed a substantial reduction in lambda GC relative to the provided PCs for all tested ancestry-trait pairs (**Supplementary note figure S1A**). The largest decline was observed for height for the AMR group, with lambda GC falling from ~1.37 to ~1.07 (**Supplementary note figure S1C**), indicating that the new PCs substantially reduce residual stratification. We use these newly recomputed PCs for all mtDNA phenotype GWAS in AoU.

**Supplementary note 5 – measures taken to avoid NUMT contamination of association results**

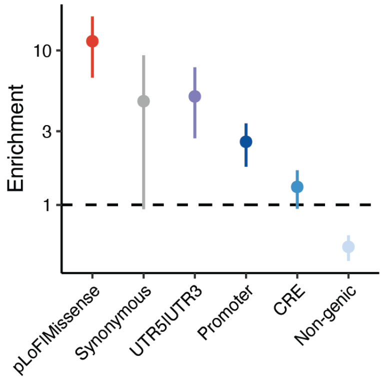


**Supplementary note figure S3**. Enrichment of annotated genomic function for variants with maximal PIP > 0.1 across all main mtDNA analyses after fine-mapping. Dotted line corresponds to no enrichment.

The study of mtDNA genome sequence variation has historically been plagued by nuclear mtDNA pseudogenes called NUMTs, which can produce spurious low-heteroplasmy variant calls due to nucDNA reads from these homologous regions mis-mapping to mtDNA. Multiple lines of evidence support the robustness of our current results:

1. Our mtSwirl pipeline maps reads to a self-reference mtDNA and known NUMT regions to reduce mis-mapping (**Supplementary figure 1**).
2. We exclude low-mtCN samples which are more prone to NUMT-derived variant calls (**Supplementary figure 7C**) (Laricchia et al., 2022).
3. We have only considered heteroplasmies >5%. Variants at lower heteroplasmy are at a higher risk of being NUMT-derived.
4. We find little-to-no evidence of paternal transmission of heteroplasmies (**Figure 4D**). Paternal transmission of mtDNA variation would be expected for NUMT-derived variants (Wei et al., 2020), however all variants for which we performed genetic analysis show strong maternal transmission in UKB.
5. Our GWAS yields numerous fine-mapped nuclear loci near genes with established roles in mtDNA biology and maintenance.
6. Associated loci tend to show small credible sets after fine-mapping (**Supplementary figures 5B, 9E**) with strong enrichment for functional variants across all main GWAS (**Supplementary note figure S3**); this would not be expected for NUMT-based associations. NUMT-based associations are expected to purely be due to read-alignment

artifact, and thus should not be correlated to variant type (e.g., missense vs. synonymous vs. noncoding).

7. Numerous sensitivity analyses yield virtually unchanged GWAS effect sizes, including correcting for overall mtCN, correcting for coverage at the mtDNA heteroplasmy site and restricting to variant calls supported by more reads than the median nucDNA coverage (**Supplementary figure 8J-8M**). We expect that NUMT-driven associations would show effect sizes correlated with coverage.
8. We successfully replicated our results in an independent cohort, finding replication even across different genetic ancestry groups (**Supplementary figure 9C, 9D**). Replication would be unlikely if recent polymorphic NUMTs were driving our results as recent polymorphic NUMTs are unlikely to appear across multiple nuclear genetic ancestry categories.
9. Finally, we collated an extensive database of 4,736 polymorphic and reference NUMT intervals using BLASTn and existing datasets (Calabrese et al., 2012; Dayama et al., 2014; Li et al., 2012; Wei et al., 2022) (**Methods**) and conservatively tested for LD $R^2 > 0.1$ between any SNP in a 20kb window around a NUMT and each genome-wide significant lead variant for our case-only heteroplasmy GWAS:

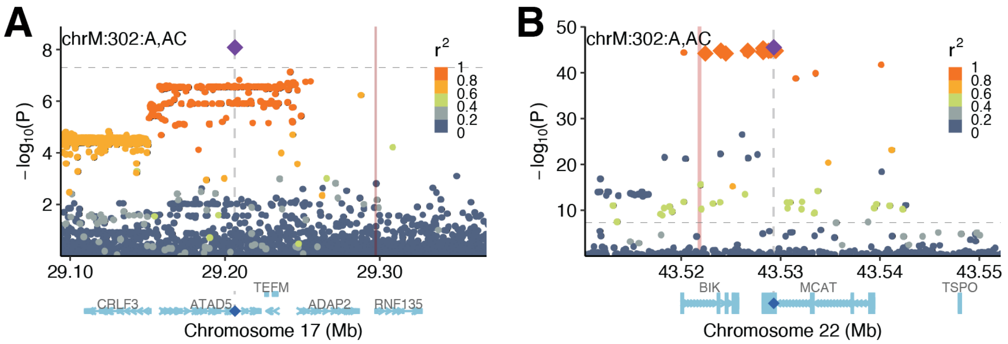


**Supplementary note figure S4**. Locuszoom plots of genetic associations between chrM:302:A,AC and **A.** the chromosome 17 locus near TEFM and **B.** the chromosome 22 locus near MCAT. Color indicates LD $R^2$ to the lead variant (purple diamond). Red ribbon corresponds to polymorphic NUMT insertion point. Large sized diamonds in panel **B** correspond to variants in the 95% credible set after fine-mapping.

Among 2,961 tested reference NUMTs, we found that only the SSBP1 locus on chromosome 7 showed LD ($R^2 > 0.9$) with a NUMT region (**Supplementary figure 9K**). Within this locus, while EUR individuals have high LD between *SSBP1* and the NUMT region, we observed a substantially shorter LD block among AFR individuals in AoU. Despite this reduced LD, GWAS in AFR individuals in AoU produced the *SSBP1*-proximal signal, suggesting that the NUMT is not driving the association (**Supplementary figure 9K**). Furthermore, the NUMT region is homologous to chrM:2740-chrM:6623; this mtDNA region does not contain chrM:302 or any of the other heteroplasmic sites found to associate with the SSBP1 locus, further reducing the likelihood of this nucDNA region confounding associations with our tested heteroplasmic sites.

Among 1,775 polymorphic NUMTs (Dayama et al., 2014; Wei et al., 2022), we find only two instances of LD between a lead SNP and any SNPs within a 20kb window around a NUMT insertion site. We note that these NUMTs would only cause spurious associations if they were in strong LD with genotyped variants, as the NUMT sequence itself is not included in the reference genome:

1. We find maximum LD $R^2$=0.606 between the TEFM locus and SNPs in a 20kb window around a polymorphic NUMT nearby. We do not believe this is likely to contaminate our signal for several reasons: (1) this is an ultra-rare (AF < 0.1%) NUMT found in the "oth" ancestry group rather than among the populations used in our UKB discovery analysis, (2) the mtDNA sequence inserted here is chrM:6866-7019, while the mtDNA heteroplasmy trait associated with the nearby locus is chrM:302, reducing the chances of reads from this locus altering the observed heteroplasmy at chrM:302, and (3) on examining local LD structure (**Supplementary note figure S4A**) it appears that there is a sharp drop-off in LD consistant with a recombination site separating our GWAS-identified locus and the true NUMT insertion point.
2. We find that the MCAT locus is within 10kb of a nearby polymorphic NUMT (**Supplementary note figure S4B**). While we cannot exclude influence of this NUMT on our GWAS signal, we believe this is not driving our signal for several reasons: (1) this locus appears significant for heteroplasmy traits, coverage discrepancy analysis, and mtCN. While the former heteroplasmy or coverage discrepancies may be sensitive to NUMT contamination, mtCN (and particularly mtCN computed using median mtDNA coverage) should be robust to polymorphic NUMT contamination; (2) the mtDNA sequence inserted is chrM:2045-2221 while the locus is associated with chrM:302, reducing the chances of reads from this locus altering the observed heteroplasmy at chrM:302; and (3) the polymorphic NUMT is ultra-rare (AF < 0.1%).

Thus, while we cannot eliminate the possibility that NUMTs may influence some of our associations, our variant calling pipeline, quality control procedure, sensitivity analysis procedure, transmission analysis, genetic analysis, fine-mapping, and LD analysis all point to our results being robust to NUMT contamination.

**Supplementary note 6 – indel variants are likely not artifacts**

Prior analyses have excluded indel variants particularly at chrM:302 for fear that these may arise due to sequencing artifact, however we believe this is unlikely to confound our results. First, we only perform genetic association testing with common indel variants (MAC > 500) in UKB. Second, it is extremely improbable that this sequencing artifact could induce associations nominating genes specifically involved in mtDNA maintenance and replication. Third, we observe that the indel variants tested show very strong quantitative maternal transmission with little evidence of paternal transmission (**Supplementary figure 7H**); this is true specifically of the length variants at chrM:302 as well (**Figure 5B**). It is very unlikely that a sequencing or PCR artifact could produce apparent quantitative maternal transmission with little detectable paternal transmission.

More generally, these variants have been used widely in population genetic and forensic studies, and several groups have directly quantified mixtures of length heteroplasmies at chrM:302 across several tissues (Lee et al., 2006; Shin et al., 2004) using size-based separation of PCR products and subsequent cloning and sequencing.

### Supplementary note 7 – Manhattan plots from UKB case-only GWAS with any genome-wide signals

We performed GWAS in UKB for 39 case-only heteroplasmy traits. To display these in a space-efficient manner, we show all clumped lead SNPs with $p < 5e-5$ in **Figure 4E** for traits that showed any genome-wide significant signal, and the same display for all tested heteroplasmies in **Supplementary figure 9A**. In this note we enclose full Manhattan plots for each of the case-only heteroplasmy GWAS in UKB which show any genome-wide significant signal – see **supplementary note figures S5, S6**.

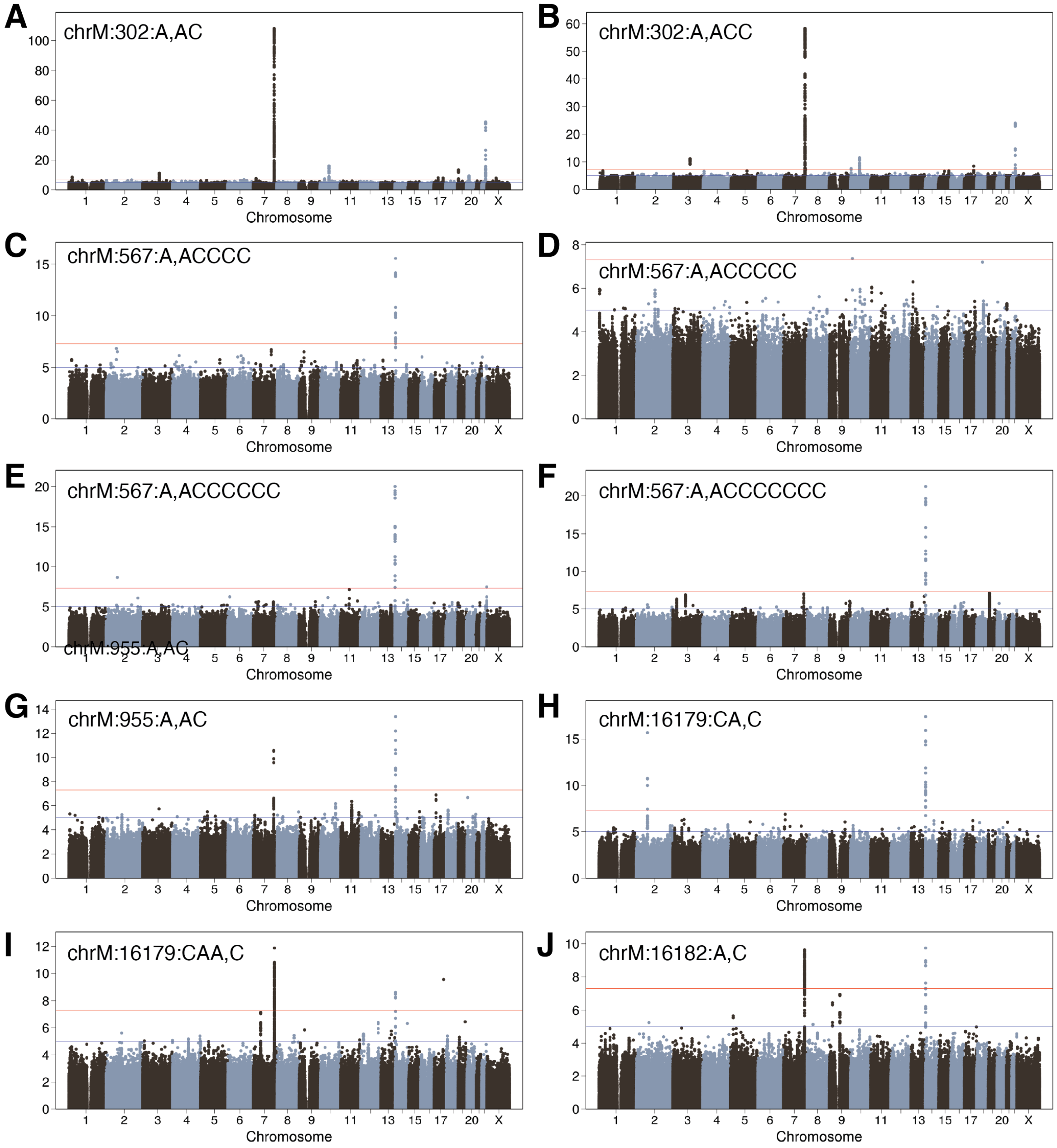


**Supplementary note figure S5**. Manhattan plots of first 10 case-only heteroplasmies with any genome-wide significant signal detected. These correspond to the lead SNPs plotted in **Figure 4E.** Red line indicates genome-wide significance, blue line indicates "suggestive". Y-axis is -log10(p-value), inset text is the case-only heteroplasmy tested.

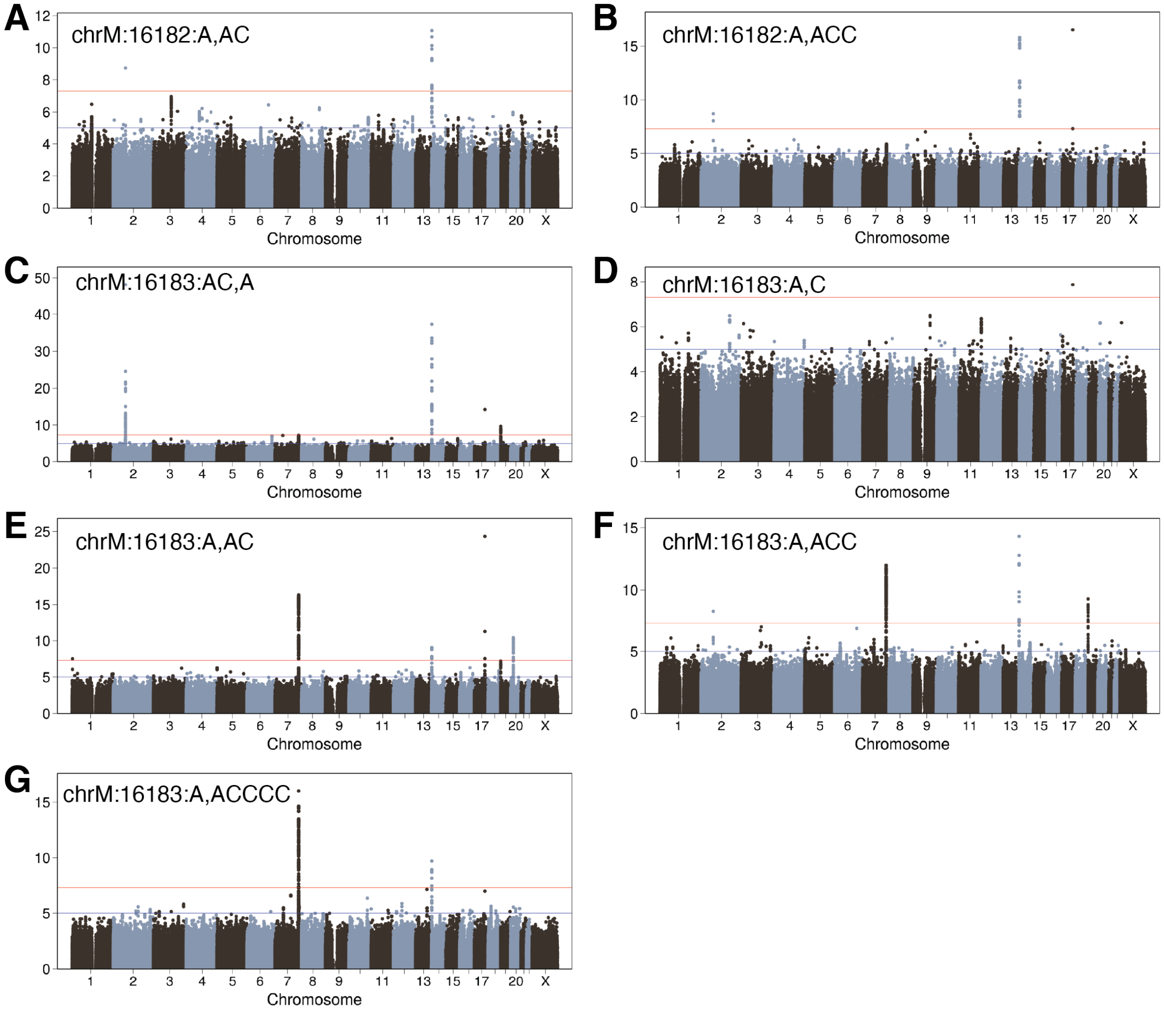


**Supplementary note figure S6**. Manhattan plots of next seven case-only heteroplasmies with any genome-wide significant signal detected. These correspond to the lead SNPs plotted in **Figure 4E.** Red line indicates genome-wide significance, blue line indicates "suggestive". Y-axis is -log10(p-value), inset text is the case-only heteroplasmy tested.
